# Pathology and genetics in a global cohort of Parkinsonian Disorders

**DOI:** 10.64898/2026.03.23.26348322

**Authors:** Lesley Y. Wu, Tessa du Toit, Tatiana Georgiades, Eleanor J. Stafford, Kristin Levine, Zih-Hua Fang, Simona Jasaityte, Ana-Luisa Gil Martinez, Patrick Cullinane, Eduardo De Pablo Fernandez, Cornelis Blauwendraat, Andrew B. Singleton, Sonja W. Scholz, Bryan J. Traynor, Nicholas Wood, John Hardy, Patrick Chinnery, Henry Houlden, Richard Cain, Claire Troakes, Viorica Chelban, Geidy E. Serrano, Djordje Gveric, Catriona McLean, Seth Love, Andrew King, Andrew C. Robinson, Federico Roncaroli, Claire Shepherd, Glenda Halliday, Laura Parkkinen, Christopher M Morris, Colin Smith, Thomas G. Beach, Steve Gentleman, Thomas T. Warner, Tammaryn Lashley, Zane Jaunmuktane, Raquel Real, Huw R Morris, Global Parkinson’s Genetic Program (GP2)

**Affiliations:** Department of Clinical and Movement Neurosciences, UCL Queen Square Institute of Neurology, London, UK; Data Tecnica International, Washington, DC, USA; The German Center for Neurodegenerative Diseases, Tübingen, Germany; Global Parkinson’s Genetics Program, Chevy Chase, MD, USA; Neurodegenerative Diseases Research Section, National Institute of Neurological Disorders and Stroke, National Institutes of Health, Bethesda, MD, USA; Department of Neurology, Johns Hopkins University School of Medicine, Baltimore, MD, USA; Neuromuscular Diseases Research Section, National Institute on Aging, National Institutes of Health, Bethesda, MD, USA; Department of Neurodegenerative Disease, UCL Queen Square Institute of Neurology, London, UK; Department of Clinical Neurosciences, University of Cambridge, Cambridge, UK; University of Bristol, Bristol, Horfield, United Kingdom; London Neurodegenerative Diseases Brain Bank, King’s College London, London, UK; Department of Pathology, Banner Sun Health Research Institute, Sun City, AZ, USA; Department of Brain Sciences, Faculty of Medicine, Imperial College London, London, UK; Victorian Brain Bank, Victoria, Australia; Geoffrey Jefferson Brain Research Centre, Division of Neuroscience, Faculty of Biology, Medicine and Health, University of Manchester, Manchester, UK; Neuroscience Research Australia, Randwick, Sydney, Australia; Department of Biomedical Science, Faculty of Health and Medicine, University of New South Wales, Sydney, Australia; School of Medical Sciences, Faculty of Medicine and Health, University of Sydney, Sydney, Australia; Department of Neuropathology and The Queen’s College, University of Oxford, Oxford, UK; Newcastle Brain Tissue Resource, NIHR Newcastle Biomedical Research Centre, Translational and Clinical Research Institute, Newcastle University, Newcastle-upon-Tyne, UK; Academic Department of Neuropathology, Institute of Neurological and Cardiovascular Research, University of Edinburgh, Edinburgh, UK; Queen Square Brain Bank for Neurological Disorders, UCL Queen Square Institute of Neurology, London, UK; Aligning Science Across Parkinson’s (ASAP) Collaborative Research Network, Chevy Chase, MD, 20815

## Abstract

**Importance:** Accurate diagnosis of neurodegenerative movement disorders is challenging because of a lack of *in vivo* biomarkers, overlapping clinical features and a delay in the emergence of pathognomonic features.

**Objective:** To evaluate clinicopathological correlation, diagnostic accuracy, genetic association with pathology, and ancestry-related differences in a multi-ancestry brain bank cohort.

**Design:** Multicentre retrospective autopsy cohort study on donors enrolled between 1985 - 2024.

**Setting:** 11 academic brain banks in the UK, US and Australia

**Participants:** Brain donors identified from participating brain banks with available brain tissue and a clinical diagnosis of Parkinson’s disease, Parkinson’s disease dementia, dementia with Lewy bodies, progressive supranuclear palsy, corticobasal syndrome, multiple system atrophy, or neurologically normal controls.

**Exposure:** Genetic variant carrier status and clinical diagnostic category.

**Main outcome:** Clinical diagnostic accuracy; Lewy body and Alzheimer’s disease pathology burden; survival; association with genetic variants and genetically inferred ancestry.

**Results:** We studied 3,353 brain donors (1281 [38.2%] female, mean [SD] age at death, 76.8 [10.6] years). Misdiagnosis rates for movement disorders ranged approximately from 10% - 20%. Clinical diagnoses of dementia with parkinsonism (PDD/DLB) were more strongly associated with Lewy body pathology than Parkinson’s disease without dementia (OR = 1·96, 95% CI = 1·30 - 3·04, p = 7·2e-04). Lewy pathology was identified in 4% of neurologically normal controls. Alzheimer’s disease co-pathology was present in 40% of cases with Lewy body disease. *GBA1* variant carriers exhibited greater Lewy body burden compared with noncarriers (OR = 1·94, 95% CI = 1·24 - 3·03, p = 0·01) or *LRRK2* carriers (OR = 7·44, 95% CI = 2·16 - 25·64, p = 0·01). Pathological diagnoses differed by ancestry, with South Asian donors more likely to have progressive supranuclear palsy pathology and Ashkenazi Jewish donors more likely to have Lewy body disease (p < 0.0001), independent of *GBA1* and *LRRK2* mutation status.

**Conclusion and Relevance:** Our findings highlight the value of integrating genetic and pathological data to improve diagnostic accuracy. The high prevalence of Alzheimer’s disease co-pathology and ancestry-related differences in pathology point to the need for biologically informed diagnostic tools. These results support the integration of genetically and pathologically stratified approaches, correlating pathology with *in vivo* biomarkers, for future therapeutic trials.

**Funding:** Medical Research Council, Global Parkinson’s Genetic Program/Aligning Science Across Parkinson’s

**Key Points:** *Question:* How do genetic variants and neuropathology influence clinical features and diagnostic accuracy in movement disorders?

*Findings:* In this multi-ancestry brain bank series including over 3000 individuals, clinical misdiagnosis was common. Dementia with parkinsonism was more strongly associated with Lewy body (LB) pathology than Parkinson’s disease without dementia, and Alzheimer’s disease co-pathology was frequent. Genetic variation was associated with pathological differences. *GBA1* carriers had greater LB burden, while *LRRK2* pathogenic variant carriers had a lower LB burden and longer survival. Pathological diagnosis differed by ancestry.

*Meaning:* Integrating genetics and neuropathology may improve diagnosis and support pathology-informed therapeutic trials.

## Introduction

Neurodegenerative movement disorders are a heterogeneous group of conditions characterised by progressive motor and cognitive impairment. Clinico-pathological studies of Parkinson’s disease (PD), dementia with Lewy bodies (DLB), progressive supranuclear palsy (PSP), corticobasal degeneration (CBD), and multiple system atrophy (MSA) have led to the development of consensus clinical diagnostic criteria, usually based on a hallmark protein-based pathology ^1–5^. Genetic factors contribute to both monogenic and complex forms of movement disorders; however, their integration into the clinico-pathological framework remains incomplete. Genetic studies are typically conducted with clinically diagnosed patients, which may overlook the impact of misdiagnosis and co-pathology. Importantly, genetic profiling can help highlight the diversity of pathological features relating to prototypic clinical presentations. For instance, variants in *GBA1* are associated with wide-spread Lewy body (LB) pathology ^6^ whereas *LRRK2* mutation carriers can present with clinically typical PD in the absence of LB at post-mortem examination ^7^. Differentiating these diseases is challenging in the early stages, as they often present overlapping clinical features, affecting the interpretation of clinical research and investigational drug studies.

Existing work integrating genetics and pathology is often restricted to small or family-based cohorts, selected by genotype, providing limited population-level insights. Movement disorders research has largely focused on individuals of European ancestry, despite growing evidence that ancestry influences disease heterogeneity, clinical presentation and outcome, including mortality ^8^.

We harmonised the genetic data and integrated with the clinical and neuropathological data from multiple brain banks, including over 3000 individuals of diverse ancestries with clinically diagnosed movement disorders and neurologically normal controls, with the aim of assessing clinico-pathological correlation in individuals carrying disease-associated and risk genetic variants and the frequency of clinical misdiagnosis. This study strengthens the knowledge base to better inform the development of future diagnostic tools and the design of clinical trials.

## Methods

### Study design and participants

We included individuals with clinically diagnosed movement disorders and neurologically unaffected controls from the Defining and Diagnosing neurodegenerative Movement Disorders through integrated analysis of Genetics and neuroPathology (MD-GAP) study (eMethods) and collaborating brain banks within the Global Parkinson’s Genetics Program (GP2). MD-GAP integrates clinical, pathological, and genetic data from autopsy-confirmed cases with neurodegenerative diseases, focusing on movement disorders. Demographic data, main clinical diagnosis, primary pathology and co-pathology were provided by brain banks (eTable 1). Clinical diagnoses included PD, PDD, DLB, PSP, Corticobasal syndrome (CBS), MSA, and Controls. Pathological diagnoses include Lewy body disease (LBD), PSP, MSA, CBD, and Other. Brains were donated between 1985 and 2024. This study followed the Strengthening the Reporting of Observational Studies in Epidemiology (STROBE) reporting guidelines for cross-sectional studies.

Ethical approval to coordinate the MD-GAP study was obtained from the UCL Research Ethics Committee (23473/001). Each contributing brain bank obtained local ethics approval for recruitment, storage, and distribution of brain donor material with written informed consent.

### Diagnostic Accuracy

The diagnostic accuracy for each pathologically defined movement disorder was evaluated by comparing the primary clinical diagnosis with neuropathology. LBDs were classified as a single pathological entity, as the distinction between PD, PDD and DLB relates to clinical features. CBS can arise from diverse underlying pathologies; we examined clinically diagnosed CBS relative to pathologically confirmed CBD.

### Neuropathological assessment and Genetic characterization

We documented amyloid-β, neurofibrillary tangle (NFT) phosphorylated tau, and α-synuclein pathology in PD, PDD, and DLB using established staging systems (McKeith ^9^, Unified staging system for LBD (USSLB) ^10^, Braak LB ^11^ and NFT stages ^12^, CERAD ^13^, and Thal ^14^ to compare pathological distributions between demented and non-demented LBD cases. For statistical power, LB staging systems were collapsed into neocortical, limbic, and brainstem LBD subtypes (eTable 2).

All cases were genotyped using the Illumina Neurobooster array (NBA) and/or underwent genome sequencing in GP2 ^15–17^. We used the PanelAPP neurodegenerative disease panel (eTables 3 and 4) to define genes of interest for neurodegenerative disease. Variants were extracted using bcftools and annotated with ANNOVAR (eMethods) and defined by ClinVar as pathogenic or likely pathogenic variants.

Genetic ancestry was determined using Genotools ^18^. Clinical sex was confirmed by genetic sex inferred from genotyping data.

### Statistical analyses

Demographic comparisons between each diagnostic and ancestry group with more than ten individuals were performed using the Kruskal-Wallis test for continuous data, followed by Bonferroni-corrected pairwise comparisons.

The accuracy of clinical diagnosis and concordance with pathological findings were evaluated with sensitivity, specificity, positive predictive value (PPV), and negative predictive value (NPV). We classified *GBA1* variants as either PD-risk or Gaucher disease-causing ^19^, and compared their frequencies between cases and controls with Fisher’s exact test. We applied proportional odds logistic regression, adjusted for sex, disease duration (DD), and age at death (AAD), with false discovery rate (FDR) correction, to assess associations between *GBA1* and *LRRK2* mutation status with LBD subtypes and Braak NFT stages, as well as the association between *APOE e4* dosage (0 - 2) and LBD subtypes. We evaluated the association between DD and *GBA1* and *LRRK2* genetic status using a Cox model, with age at onset (AAO) as covariate. We compared *MAPT* haplotype distribution across pathological diagnostic groups using the Chi-square test with FDR-corrected pairwise comparisons. We compared genetic ancestry and pathological diagnosis using Pearson’s Chi-square test.

We used R statistical software version 4·3·1.

## Results

We identified 3,353 individuals with antemortem primary clinical diagnoses of a movement disorder or controls: 1171 PD, 399 PDD, 227 DLB, 811 Parkinson’s Plus syndromes (PPS; 491 PSP, 244 MSA, 76 CBS) and 745 neurologically normal controls (Table 1).

**Table 1:**
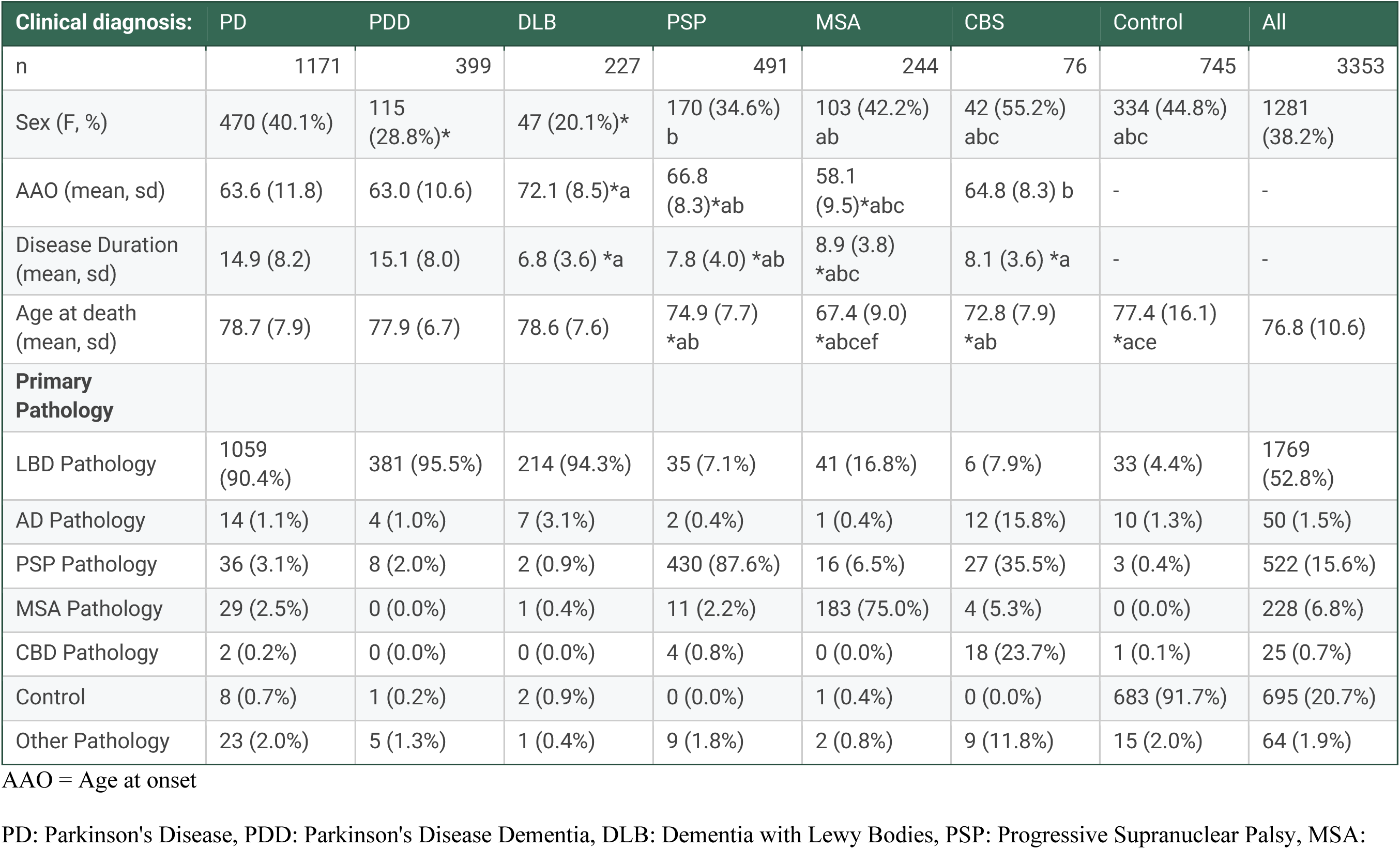
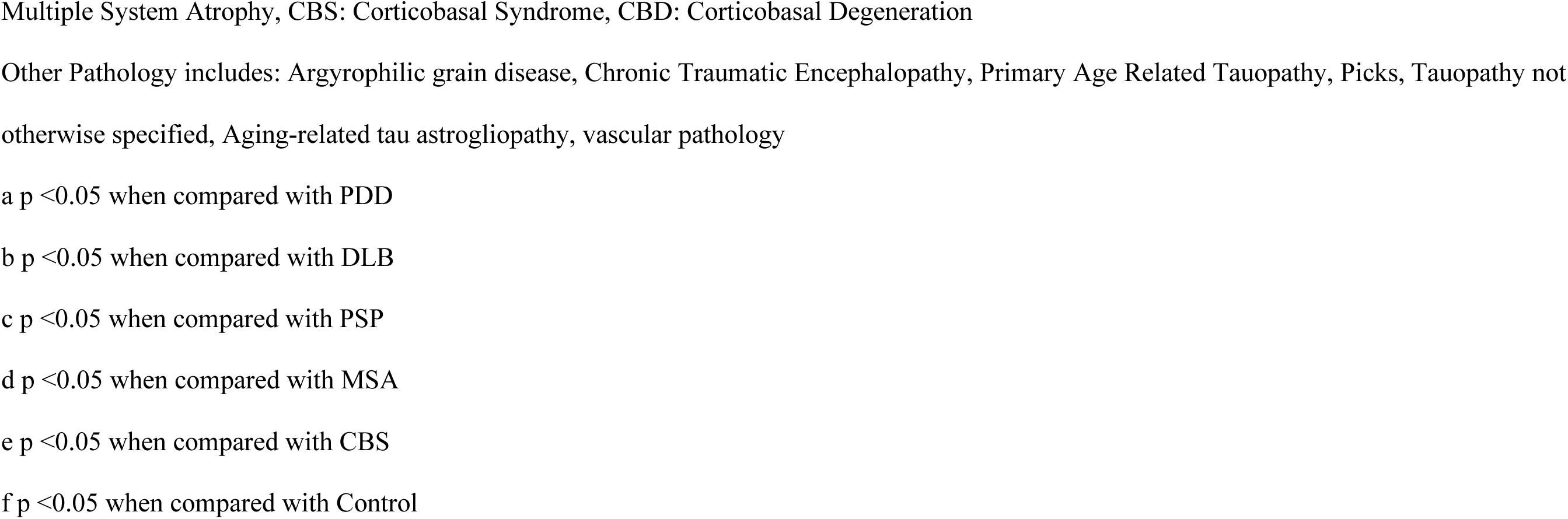
Demographic features and underlying primary pathology in 3403 clinically diagnosed movement disorder cases and neurologically healthy controls.

| Clinical diagnosis: | PD | PDD | DLB | PSP | MSA | CBS | Control | All |
| --- | --- | --- | --- | --- | --- | --- | --- | --- |
| n | 1171 | 399 | 227 | 491 | 244 | 76 | 745 | 3353 |
| Sex (F, %) | 470 (40.1%) | 115 (28.8%)* | 47 (20.1%)* | 170 (34.6%)<br>b | 103 (42.2%)<br>ab | 42 (55.2%)<br>abc | 334 (44.8%)<br>abc | 1281 (38.2%) |
| AAO (mean, sd) | 63.6 (11.8) | 63.0 (10.6) | 72.1 (8.5)*a | 66.8 (8.3)*ab | 58.1 (9.5)*abc | 64.8 (8.3) b | - | - |
| Disease Duration (mean, sd) | 14.9 (8.2) | 15.1 (8.0) | 6.8 (3.6) *a | 7.8 (4.0) *ab | 8.9 (3.8) *abc | 8.1 (3.6) *a | - | - |
| Age at death (mean, sd) | 78.7 (7.9) | 77.9 (6.7) | 78.6 (7.6) | 74.9 (7.7) *ab | 67.4 (9.0) *abcef | 72.8 (7.9) *ab | 77.4 (16.1) *ace | 76.8 (10.6) |
| **Primary Pathology** |  |  |  |  |  |  |  |  |
| LBD Pathology | 1059 (90.4%) | 381 (95.5%) | 214 (94.3%) | 35 (7.1%) | 41 (16.8%) | 6 (7.9%) | 33 (4.4%) | 1769 (52.8%) |
| AD Pathology | 14 (1.1%) | 4 (1.0%) | 7 (3.1%) | 2 (0.4%) | 1 (0.4%) | 12 (15.8%) | 10 (1.3%) | 50 (1.5%) |
| PSP Pathology | 36 (3.1%) | 8 (2.0%) | 2 (0.9%) | 430 (87.6%) | 16 (6.5%) | 27 (35.5%) | 3 (0.4%) | 522 (15.6%) |
| MSA Pathology | 29 (2.5%) | 0 (0.0%) | 1 (0.4%) | 11 (2.2%) | 183 (75.0%) | 4 (5.3%) | 0 (0.0%) | 228 (6.8%) |
| CBD Pathology | 2 (0.2%) | 0 (0.0%) | 0 (0.0%) | 4 (0.8%) | 0 (0.0%) | 18 (23.7%) | 1 (0.1%) | 25 (0.7%) |
| Control | 8 (0.7%) | 1 (0.2%) | 2 (0.9%) | 0 (0.0%) | 1 (0.4%) | 0 (0.0%) | 683 (91.7%) | 695 (20.7%) |
| Other Pathology | 23 (2.0%) | 5 (1.3%) | 1 (0.4%) | 9 (1.8%) | 2 (0.8%) | 9 (11.8%) | 15 (2.0%) | 64 (1.9%) |

AAO = Age at onset
PD: Parkinson's Disease, PDD: Parkinson's Disease Dementia, DLB: Dementia with Lewy Bodies, PSP: Progressive Supranuclear Palsy, MSA:
Multiple System Atrophy, CBS: Corticobasal Syndrome, CBD: Corticobasal Degeneration
Other Pathology includes: Argyrophilic grain disease, Chronic Traumatic Encephalopathy, Primary Age Related Tauopathy, Picks, Tauopathy not
otherwise specified, Aging-related tau astrogliopathy, vascular pathology
a $p < 0.05$ when compared with PDD
b $p < 0.05$ when compared with DLB
c $p < 0.05$ when compared with PSP
d $p < 0.05$ when compared with MSA
e $p < 0.05$ when compared with CBS
f $p < 0.05$ when compared with Control

### Demographics

AAO differed significantly across clinical diagnostic groups. DLB cases had a later disease onset than all other groups but a shorter disease duration compared to PD and PDD, suggesting a more aggressive disease course (p < 0·05, Table 1). MSA cases had a significantly earlier onset and died at a younger age compared to other groups (p < 0·05). Overall, PPS were associated with shorter disease duration compared with PD and PDD (p < 0·05). Individuals of South Asian (SAS) ancestry died at a significantly younger age than individuals of European or Ashkenazi Jewish (AJ) ancestry (p < 0·001, eTable 5).

### Diagnostic accuracy across movement disorders

This study confirms the high rate of clinical misdiagnosis when compared to the pathological gold standard ^20^, with PPV varying across disease groups (Table 2). A clinical diagnosis of PD/PDD/DLB was predictive of underlying LBD pathology (PPV 92·0%, 95% CI = 90·7 - 93·2%). The presence of dementia significantly increased diagnostic accuracy: PDD/DLB was nearly twice as likely to correspond to LBD pathology compared to PD (OR = 1·96, 95% CI = 1·30 - 3·04, p = 7·2e-04). Conversely, PD without dementia showed lower diagnostic accuracy (PPV 90·4%). CBS exhibited the lowest PPV at 23·7%, reflecting the heterogeneous nature of the disease and the challenge of accurately predicting CBD pathology. MSA and CBS both had high specificity (98·1% and 98·3% respectively), while specificity for clinically diagnosed PD/PDD/DLB was the lowest (91·9%), indicating misdiagnosis of other movement disorders as PD/PDD/DLB in almost 8·0% of cases.

**Table 2.**
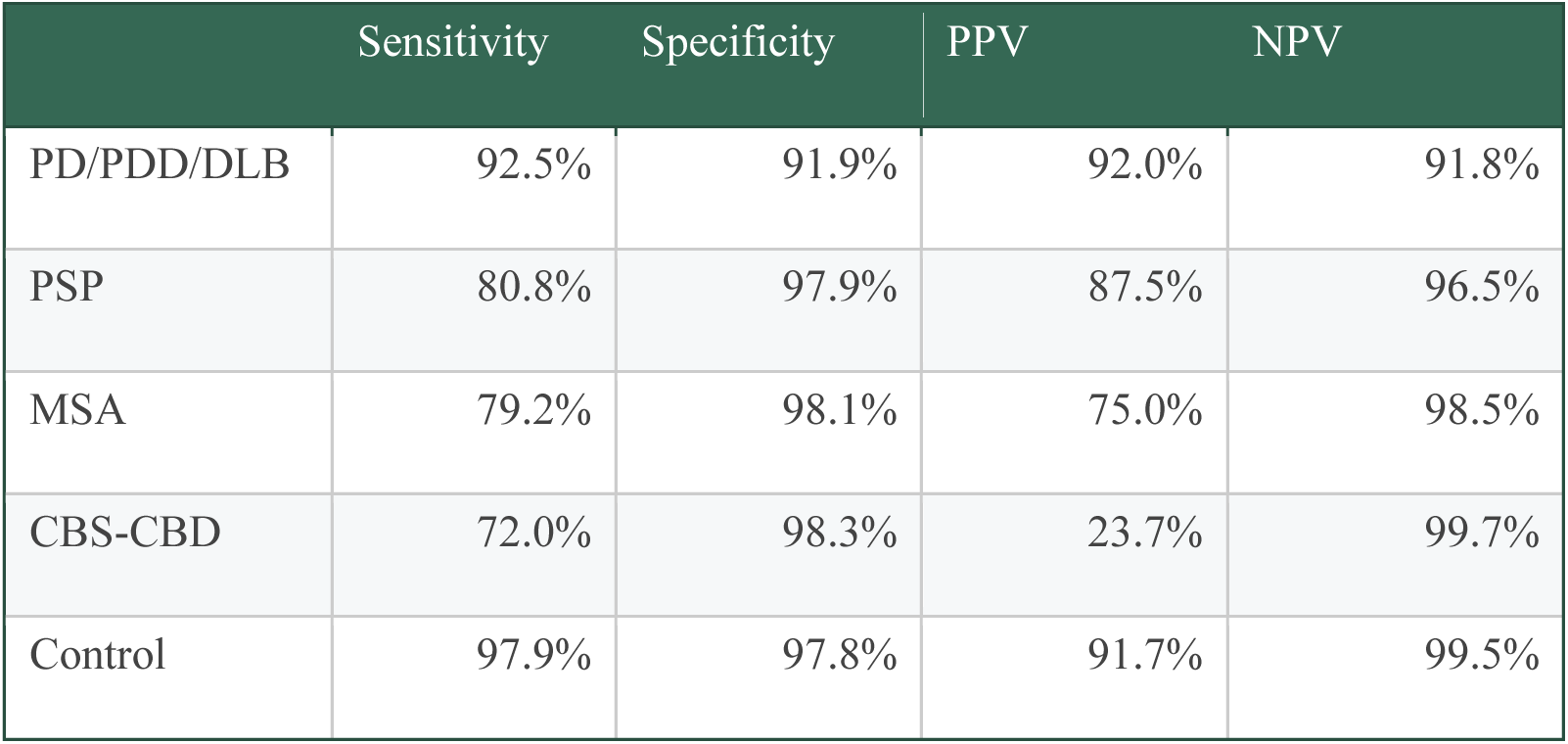
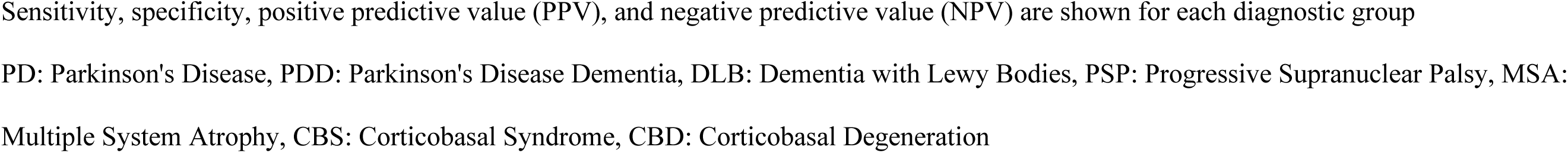
Diagnostic accuracy across clinical movement disorder diagnoses compared to neuropathological confirmation.

There was overlap between PSP and PD: 36/1171 (3·1%) clinically diagnosed PD patients had primary PSP pathology and 35/491 (7·1%) clinically diagnosed PSP patients had primary LBD pathology. Among clinically diagnosed MSA cases, 41/244 (16·8%) had primary LBD and 16/244 (6·5%) had PSP pathology. Clinically diagnosed CBS was often due to PSP (27/76; 35·5%) or Alzheimer’s disease (AD) pathology (12/76; 15·8%) at autopsy. Background neurodegenerative pathology was identified in 62/745 (8·3%) clinical controls, including 80/745 (10·7%) individuals who died before age 65. LB pathology was present in 33/745 (4·4%) clinical controls, including neocortical involvement in 10/33 (30·3%), of whom 2/10 (20·0 %) carried an *APOE* ε4 allele (eTable 6).

Co-pathology was present in 1102/1312 (84·0%) of clinically affected cases with available data. AD pathology was most frequent, present in 426/1064 (40·0%) LBD cases (eTable 7) and associated with more extensive LB distribution (OR = 2·61, 95% CI = 1·79 - 3·80, p = 5·8e-07). AD co-pathology was less common in PSP and CBD (35/216; 16·2 % and 2/12; 16·7%, respectively), which more often had additional tau pathology or other co-pathologies. Across diagnostic groups, co-pathologies was associated with later AAO (β=1·66, 95% CI = 0·99 - 2·32, p = 1·1e-06), as compared with patients reported to have no co-pathology.

### Dementia in LBD

We examined the association between dementia (defined by the clinical diagnosis of PDD and DLB) and pathological staging for LB, NFT, and amyloid-β plaques in LBD. A stepwise increase in the severity of both LB and AD pathology was observed across the clinical spectrum. Greater burdens of LB and AD pathology were independently associated with a clinical diagnosis of dementia in individuals with PDD and DLB compared with PD without dementia. Neocortical LB pathology was present in 113/140 (80·7%) of individuals with DLB, compared with 194/288 (67·4%) in PDD and 396/744 (53·2%) in PD (X^2^ = 45.75, df = 3, p = 1.16e^-10^). Similar gradients were observed in Braak NFT, CERAD, and Thal phase, supporting a cumulative increased burden of AD and LBD pathology from PD to PDD to DLB (eTable 8).

### Genetic variation

We assessed the frequency of individuals carrying common and rare variants in genes previously associated with movement disorders (Table 3). Individuals with pathologically defined LBD were more likely than pathological controls to carry *GBA1* GD-causing variants (OR = 5·65, 95% CI=1·35 - 23·66, p = 5e-03), and *GBA1* PD risk variants (OR = 1·58, 95% CI=1·07 - 2·34, p = 0·02) (eTable 9). No significant differences in *GBA1* variant frequency were observed in patients with primary PSP, MSA, or CBD pathology compared to controls. LBD cases carrying *GBA1* variants (regardless of clinical diagnosis) exhibited significantly more widespread LB pathology compared to LBD cases without any known mutations (OR = 1·94, 95% CI = 1·24 - 3·03, p = 0·01), or with an *LRRK2* mutation (OR = 7·44, 95% CI = 2·16 - 25·64, p = 0·01), after adjusting for DD (Figure 1).

**Table 3.**
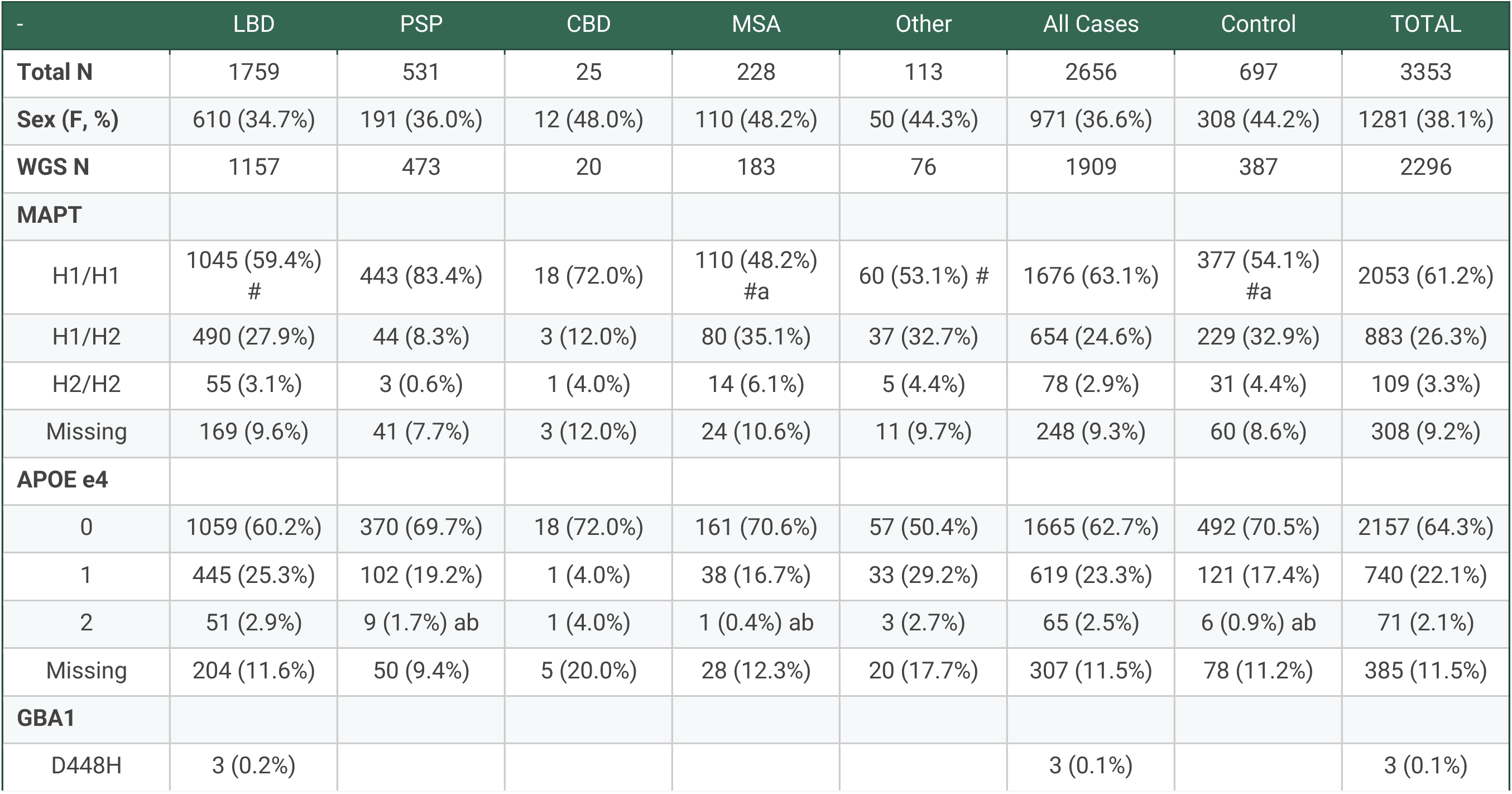
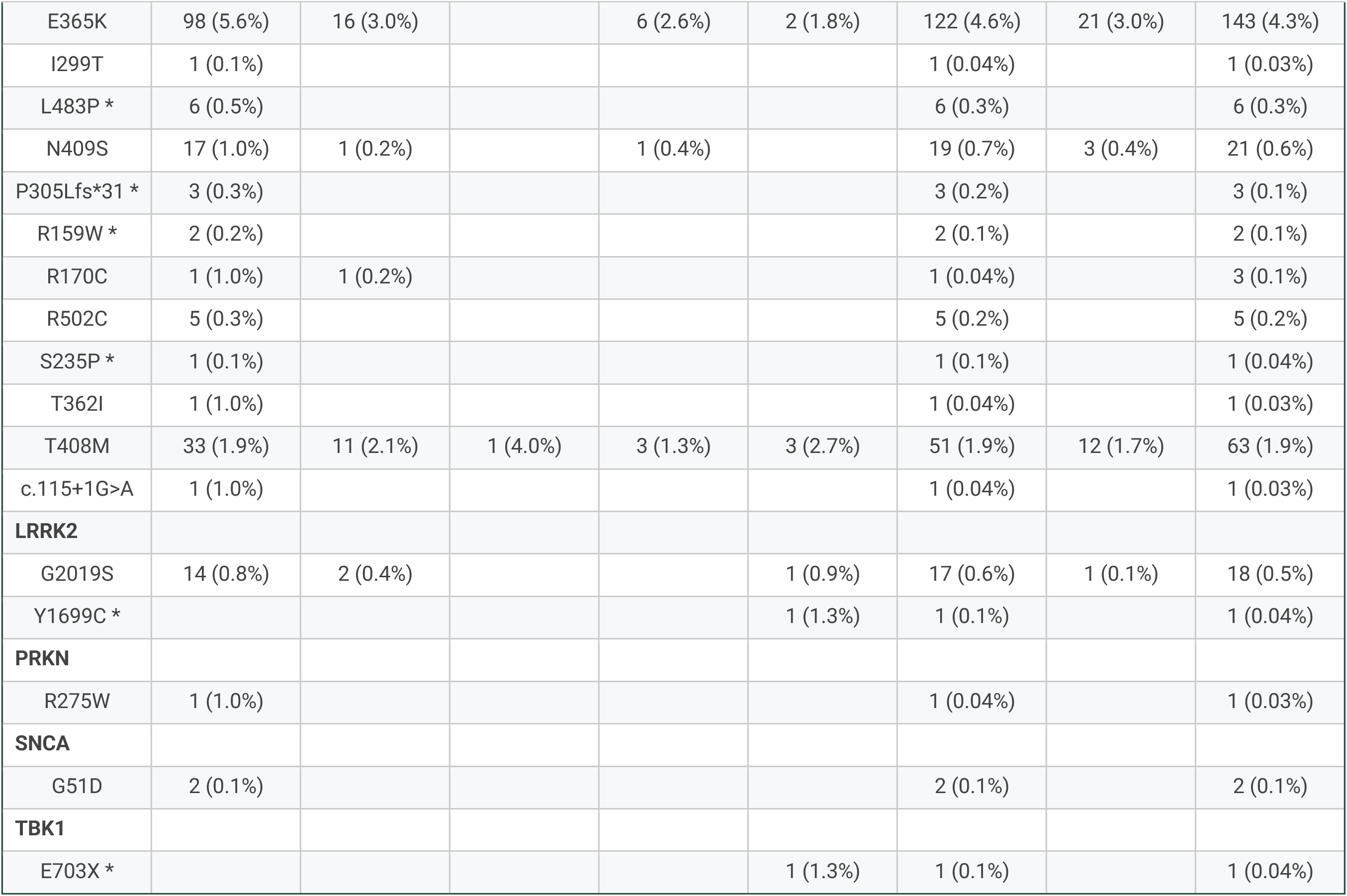
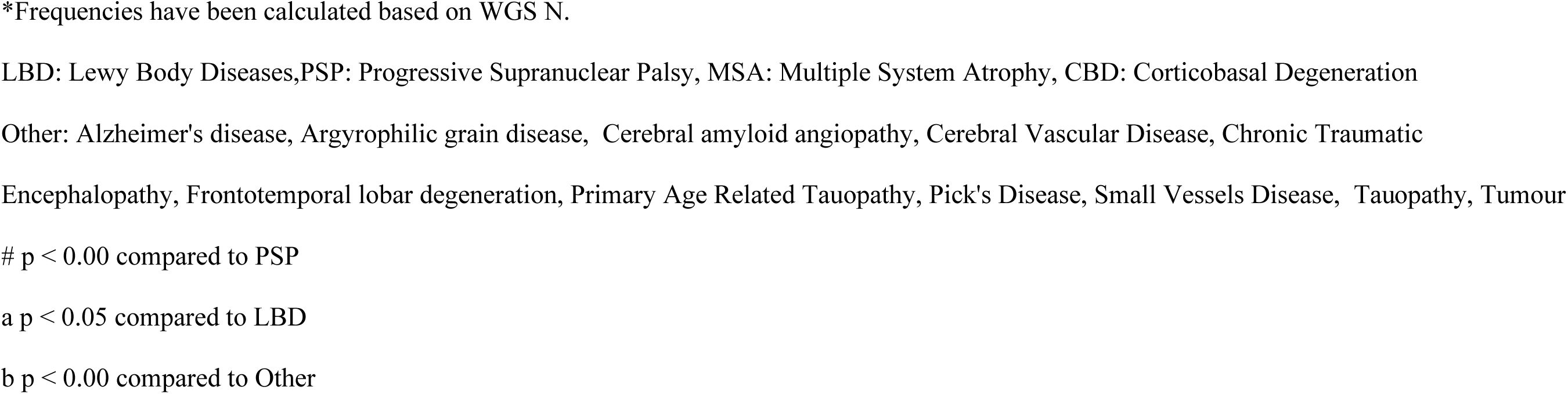
Genetic variation across pathologically defined diagnostic groups.

| - | LBD | PSP | CBD | MSA | Other | All Cases | Control | TOTAL |
| --- | --- | --- | --- | --- | --- | --- | --- | --- |
| **Total N** | 1759 | 531 | 25 | 228 | 113 | 2656 | 697 | 3353 |
| **Sex (F, %)** | 610 (34.7%) | 191 (36.0%) | 12 (48.0%) | 110 (48.2%) | 50 (44.3%) | 971 (36.6%) | 308 (44.2%) | 1281 (38.1%) |
| **WGS N** | 1157 | 473 | 20 | 183 | 76 | 1909 | 387 | 2296 |
| **MAPT** |  |  |  |  |  |  |  |  |
| H1/H1 | 1045 (59.4%)<br># | 443 (83.4%) | 18 (72.0%) | 110 (48.2%)<br>#a | 60 (53.1%) # | 1676 (63.1%) | 377 (54.1%)<br>#a | 2053 (61.2%) |
| H1/H2 | 490 (27.9%) | 44 (8.3%) | 3 (12.0%) | 80 (35.1%) | 37 (32.7%) | 654 (24.6%) | 229 (32.9%) | 883 (26.3%) |
| H2/H2 | 55 (3.1%) | 3 (0.6%) | 1 (4.0%) | 14 (6.1%) | 5 (4.4%) | 78 (2.9%) | 31 (4.4%) | 109 (3.3%) |
| Missing | 169 (9.6%) | 41 (7.7%) | 3 (12.0%) | 24 (10.6%) | 11 (9.7%) | 248 (9.3%) | 60 (8.6%) | 308 (9.2%) |
| **APOE e4** |  |  |  |  |  |  |  |  |
| 0 | 1059 (60.2%) | 370 (69.7%) | 18 (72.0%) | 161 (70.6%) | 57 (50.4%) | 1665 (62.7%) | 492 (70.5%) | 2157 (64.3%) |
| 1 | 445 (25.3%) | 102 (19.2%) | 1 (4.0%) | 38 (16.7%) | 33 (29.2%) | 619 (23.3%) | 121 (17.4%) | 740 (22.1%) |
| 2 | 51 (2.9%) | 9 (1.7%) ab | 1 (4.0%) | 1 (0.4%) ab | 3 (2.7%) | 65 (2.5%) | 6 (0.9%) ab | 71 (2.1%) |
| Missing | 204 (11.6%) | 50 (9.4%) | 5 (20.0%) | 28 (12.3%) | 20 (17.7%) | 307 (11.5%) | 78 (11.2%) | 385 (11.5%) |
| **GBA1** |  |  |  |  |  |  |  |  |
| D448H | 3 (0.2%) |  |  |  |  | 3 (0.1%) |  | 3 (0.1%) |
| E365K | 98 (5.6%) | 16 (3.0%) |  | 6 (2.6%) | 2 (1.8%) | 122 (4.6%) | 21 (3.0%) | 143 (4.3%) |
| I299T | 1 (0.1%) |  |  |  |  | 1 (0.04%) |  | 1 (0.03%) |
| L483P * | 6 (0.5%) |  |  |  |  | 6 (0.3%) |  | 6 (0.3%) |
| N409S | 17 (1.0%) | 1 (0.2%) |  | 1 (0.4%) |  | 19 (0.7%) | 3 (0.4%) | 21 (0.6%) |
| P305Lfs*31 * | 3 (0.3%) |  |  |  |  | 3 (0.2%) |  | 3 (0.1%) |
| R159W * | 2 (0.2%) |  |  |  |  | 2 (0.1%) |  | 2 (0.1%) |
| R170C | 1 (1.0%) | 1 (0.2%) |  |  |  | 1 (0.04%) |  | 3 (0.1%) |
| R502C | 5 (0.3%) |  |  |  |  | 5 (0.2%) |  | 5 (0.2%) |
| S235P * | 1 (0.1%) |  |  |  |  | 1 (0.1%) |  | 1 (0.04%) |
| T362I | 1 (1.0%) |  |  |  |  | 1 (0.04%) |  | 1 (0.03%) |
| T408M | 33 (1.9%) | 11 (2.1%) | 1 (4.0%) | 3 (1.3%) | 3 (2.7%) | 51 (1.9%) | 12 (1.7%) | 63 (1.9%) |
| c.115+1G>A | 1 (1.0%) |  |  |  |  | 1 (0.04%) |  | 1 (0.03%) |
| **LRRK2** |  |  |  |  |  |  |  |  |
| G2019S | 14 (0.8%) | 2 (0.4%) |  |  | 1 (0.9%) | 17 (0.6%) | 1 (0.1%) | 18 (0.5%) |
| Y1699C * |  |  |  |  | 1 (1.3%) | 1 (0.1%) |  | 1 (0.04%) |
| **PRKN** |  |  |  |  |  |  |  |  |
| R275W | 1 (1.0%) |  |  |  |  | 1 (0.04%) |  | 1 (0.03%) |
| **SNCA** |  |  |  |  |  |  |  |  |
| G51D | 2 (0.1%) |  |  |  |  | 2 (0.1%) |  | 2 (0.1%) |
| **TBK1** |  |  |  |  |  |  |  |  |
| E703X * |  |  |  |  | 1 (1.3%) | 1 (0.1%) |  | 1 (0.04%) |

\*Frequencies have been calculated based on WGS N.
LBD: Lewy Body Diseases, PSP: Progressive Supranuclear Palsy, MSA: Multiple System Atrophy, CBD: Corticobasal Degeneration
Other: Alzheimer's disease, Argrophilic grain disease, Cerebral amyloid angiopathy, Cerebral Vascular Disease, Chronic Traumatic
Encephalopathy, Frontotemporal lobar degeneration, Primary Age Related Tauopathy, Pick's Disease, Small Vessels Disease, Tauopathy, Tumour

**Figure 1.**
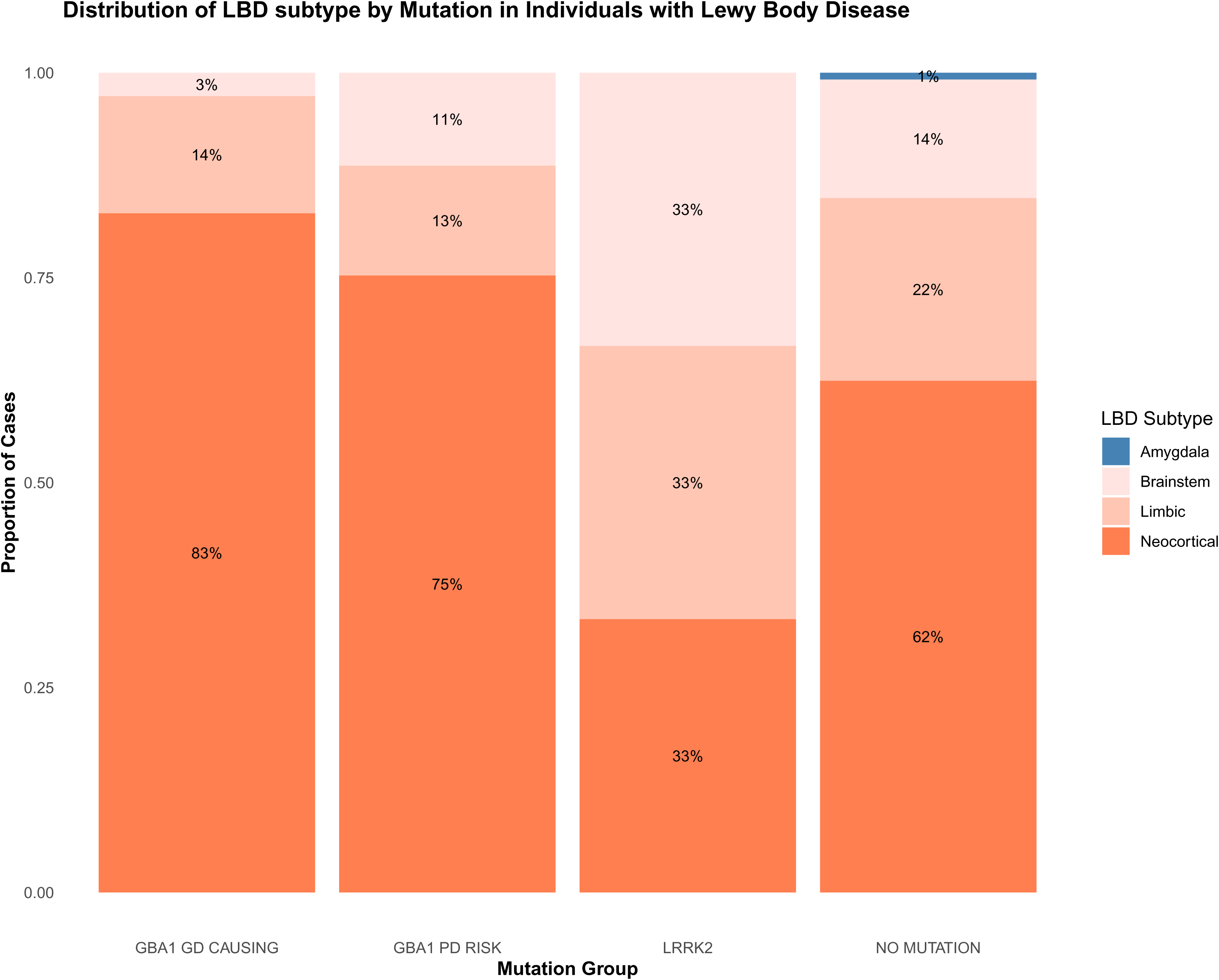
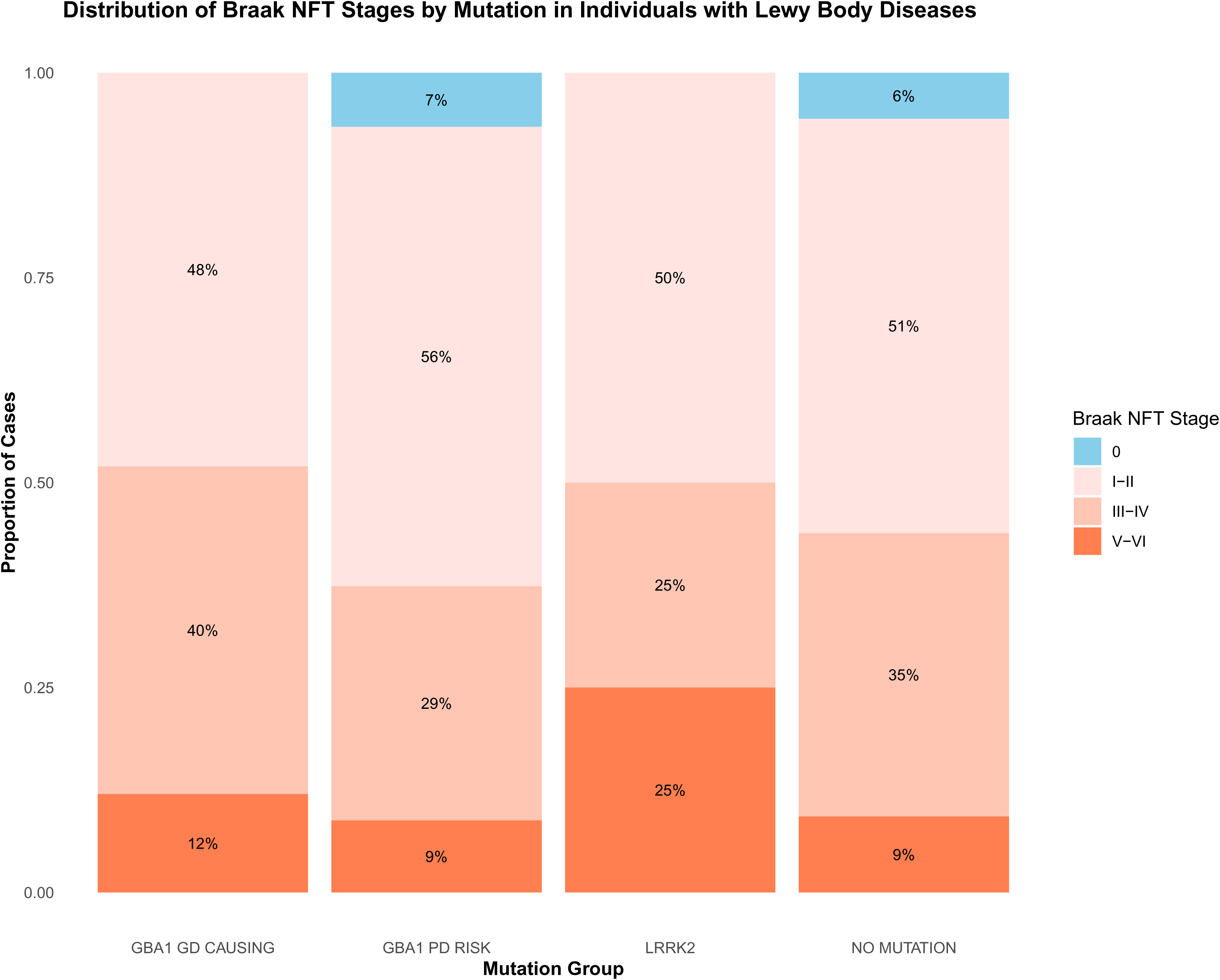
Distribution of LBD subtype and Braak NFT stages by mutation group in individuals with LBD. Stacked bar plots illustrate the proportion of pathologically diagnosed Lewy body disease (LBD) cases within each mutation group stratified by (A) LBD subtypes and (B) Braak neurofibrillary tangle (NFT). Mutation groups include *GBA1* Gaucher disease (GD)-causing mutations (R159W, R170C, S235P, I299T, P305Lfs*31, T362, N409S, D448H, L483P, R502C), *GBA1* Parkinson’s disease (PD) risk variants (E365K, T408M), *LRRK2* variants (G2019S and Y1699C) and cases without pathogenic mutations. Panel A shows that individuals carrying *GBA1* mutations, particularly GD-causing variants, are more likely to have a higher LBD pathology burden. Panel B indicates a relatively even distribution of Braak NFT stages across groups, with a trend toward higher NFT stages in *LRRK2* carriers.

We identified 19 individuals with clinically diagnosed movement disorders who carried *LRRK2* mutations (eTable 10). Of these, 18 had the p.G2019S mutation, and one carried the p.Y1699C mutation. AJ ancestry was present in 4/19 (21·0%) *LRRK2* cases versus 59/3384 (1·7%) in the remainder of the cohort, indicating significant enrichment of *LRRK2* among individuals of AJ descent. At a pathological level, LB pathology was found in 13/19 (68·4%) individuals, while the remaining cases exhibited PSP, FTLD, Other, or no pathology. *LRRK2* pathogenic variant carriers were less likely to exhibit advanced stages of LB pathology compared to individuals without mutations (OR = 0·26, 95% CI = 0·08 - 0·84, p = 0·05). Conversely, these individuals showed a trend toward higher Braak NFT stages; however, this was not statistically significant, (Figure 1), and NFT stage distributions were similar to age-matched controls. In the survival analysis (Figure 2), *LRRK2* variant carriers showed a significantly reduced hazard, indicating longer survival compared to individuals without mutations (HR = 0·60, 95% CI = 0·38 - 0·95, p = 0·02) and those carrying *GBA1* PD risk mutations (HR = 0·58, 95% CI = 0·36 - 0·94, p = 0·03). In contrast, *GBA1* variants (GD-causing or PD risk, and combined) carriers did not differ significantly from the reference group of LBD disease patients without *GBA1* variants (HR = 0·99, 95% CI = 0·85 - 1·15, p=0·88).

**Figure 2.**
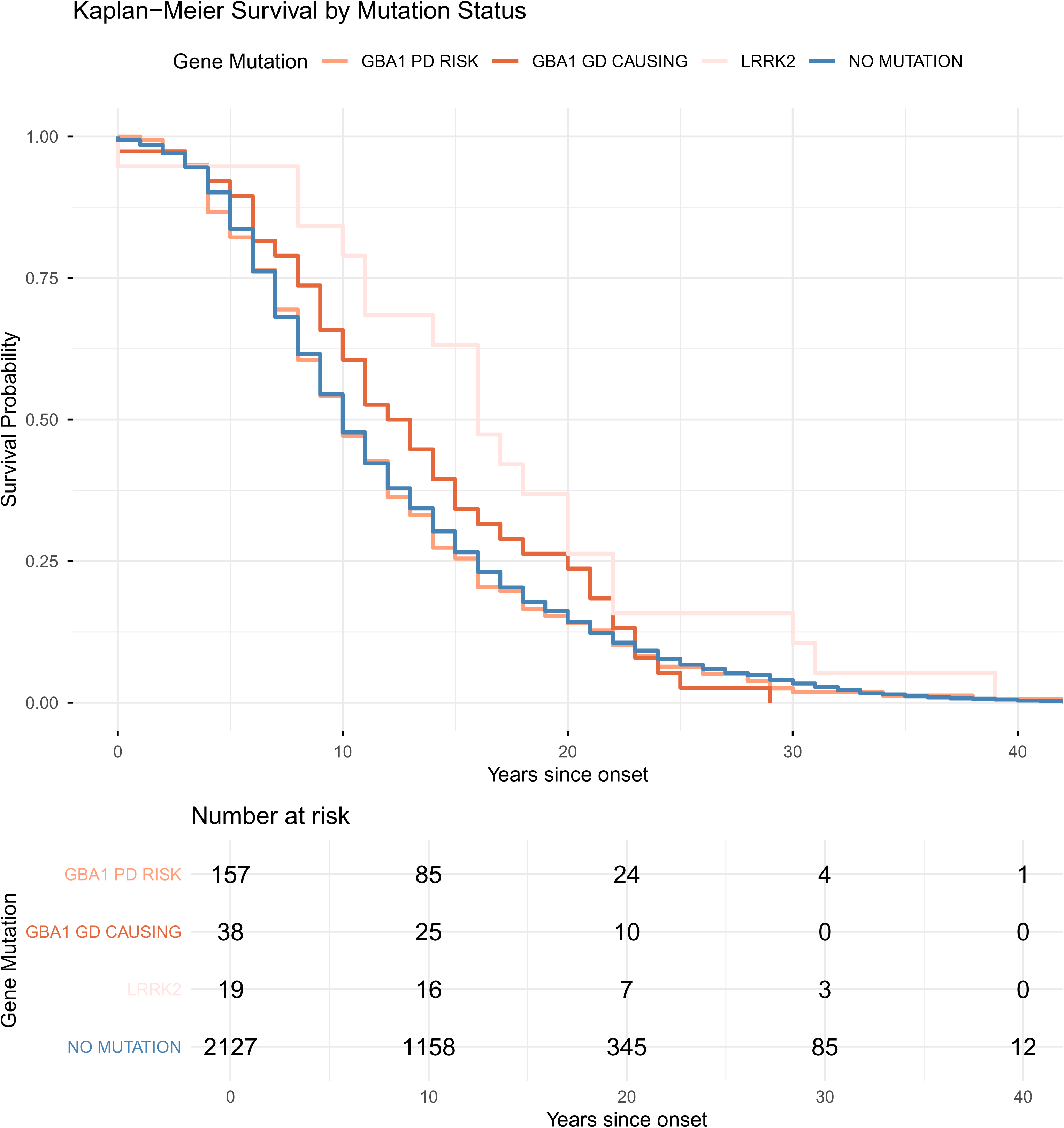
Kaplan-Meier (KM) survival curve in individuals with *GBA1*, *LRRK2* and without known mutations. KM survival curves showing disease duration (years from symptom onset to death) stratified by mutation group. Mutation groups include *GBA1* Gaucher disease (GD)-causing variants (R159W, R170C, S235P, I299T, P305Lfs*31, T362, N409S, D448H, L483P, R502C), *GBA1* PD risk variants (E365K, T408M), *LRRK2* variants (G2019S and Y1699C), and cases without pathogenic mutations. *LRRK2* carriers had a significantly longer disease duration compared to individuals without mutations (HR = 0.60, 95% CI = 0.38 - 0.95, p = 0.02) or those carrying *GBA1* PD risk mutations (HR = 0.58, 95% CI = 0.36 - 0.94, p = 0.03).

In addition to *GBA1* and *LRRK2*, pathogenic variants were identified in *SNCA, PRKN,* and *TBK1* genes. The pathogenic *TBK1* variant was identified in a case of clinical CBS with Primary Progressive Aphasia, with FTLD-TDP43 type A neuropathology, as previously reported ^21^.

*MAPT* haplotype frequencies differed significantly across pathological diagnostic groups. H1/H1 is most frequent in PSP, with significantly higher proportions compared to all other groups except CBD, and the least frequent in MSA (X^2^ = 160, df = 5, p = 8·2e-33, Table 3).

*APOE ε*4 dosage differed significantly between groups (X^2^ = 54·5, df = 5, p = 1.6e-10). LBD cases had the highest burden of ε4 dosage compared to MSA, PSP, and Controls (Table 3). In pathologically confirmed LBD cases, each additional copy of the *APOE* ε4 allele was associated with a two-fold increase in the odds of having more widespread LB pathology (OR = 2·25, 95% CI = 1·75 - 2·90, p = 4·2e-10), independent of age, sex, and brain bank.

### Ancestry analysis

We found a significant association between ancestry and pathological diagnosis (X^2^ = 35·5, df =2, p= 1·95e-08, eTable 5). LBD was more common in individuals of AJ ancestry compared to the SAS population, whereas PSP was more frequent in SAS (eFigure 1). The association between AJ ancestry and LBD remained following removal of *LRRK2*, *GBA1* risk, and rare variant carriers.

## Discussion

We have completed a large multi-center autopsy-confirmed analysis integrating clinical, genetic, and pathological data in neurodegenerative movement disorders. Our findings reinforce the complexity of clinico-pathological correlations in LBD and PPS and highlight the need for *in vivo* biomarkers for identifying underlying pathology.

We assessed misdiagnosis of Parkinsonian syndromes by comparing primary clinical diagnosis to post-mortem diagnosis. Despite our cohort being over 30 times larger than that of Hughes *et al.* (1992) ^22^, clinical misdiagnosis rates remained similar (10-20%), consistent with other clinicopathological studies ^20^ and longitudinal cohorts such as CamPaIGN ^23^. Diagnostic discordance likely reflects limitations of clinical criteria, particularly in early disease ^24,25^, and may be further influenced by copathology, underscoring the need for careful documentation of mixed pathologies in future clinicopathological studies. We showed that clinical diagnostic accuracy for LBD increases in the presence of dementia, consistent with previous research showing that symptoms such as visual hallucinations strongly support underlying LB pathology, and occur less commonly in PSP or MSA ^26^. This study highlights a high rate of misdiagnosis in MSA, where over 20% of clinically diagnosed cases had alternative pathology at autopsy, most commonly LBD or PSP. While some misclassification may reflect limited familiarity with the clinical features of PPS, these findings underscore the inherent difficulty in differentiating movement disorders, particularly in the early stages. Recent advances in seed amplification assays (SAAs) and emerging protein-based imaging modalities may help define underlying pathology during life. α-synuclein seeding activity has been detected in PSP and CBS using CSF α-synuclein SAAs, and in PSP may be associated with differences in clinical disease course ^27^. These findings align with our autopsy data showing synuclein co-pathology in a subset of PPS. The increasing focus on biological disease classification and *in vivo* biomarkers has renewed interest in grouping PD, PDD, and DLB as a unified disease entity characterised by neuronal synuclein pathology ^28,29^. Although these disorders may be indistinguishable at the individual pathological level^30^, group-level differences are evident. Previous studies, confirmed by our cohort, have shown that dementia in PD/DLB is strongly associated with a higher burden of cortical LB pathology ^31^. Differences between PDD and DLB are also apparent, with DLB, defined by primary or early dementia, associated with a later age of onset and a higher rate of AD pathology^32^, as observed in our cohort. The presence of multiple pathologies was associated with older age at onset, possibly reflecting an age-related decline in protein clearance mechanisms and the accumulation of pathological proteins ^33^.

Large-scale genotyping and genome sequencing allow the rapid definition of relevant common and rare genetic variations. We identified pathogenic/likely pathogenic variants in five genes previously implicated in neurodegenerative movement disorders. The most frequently observed mutations were in *GBA1* and *LRRK2*, consistent with their established role in parkinsonism. The frequencies of *GBA1* GD-causing, PD risk, and *LRRK2* variants in our autopsy cohort are comparable to those reported in living UK cohorts ^34,35^. *GBA1* variant carriers exhibited a broader distribution of LB pathology, contrasting with smaller prior studies showing no significant differences between carriers and non-carriers ^36,37^. These discrepancies may reflect limited statistical power in earlier studies, and highlights the need for large-scale genetic-pathological studies.

In LBD, *LRRK2* variant carriers had longer disease duration than *GBA1* carriers and non-carriers, consistent with previous reports of a milder *LRRK2*-associated disease course^38^. All *LRRK2* carriers (n = 19) exhibited some degree of NFT pathology, including two p.G2019S carriers clinically diagnosed with PD but pathologically confirmed as PSP, consistent with previous reports describing PSP-like tau pathology in *LRRK2* p.G2019S carriers ^39,40^. Interestingly, one clinically diagnosed *LRRK2* PD patient did not have pathology at autopsy. Although α-synuclein oligomer levels were not assessed, emerging evidence suggests *LRRK2*-PD without LB pathology may involve higher levels of α-synuclein oligomers in the brain^41,42^. These findings highlight the pleiotropic and heterogeneous pathological effects of pathogenic *LRRK2* variants.

Sample sizes for non-European ancestry groups remained limited, precluding ancestry-specific analyses. SAS ancestry was more frequently associated with PSP and AJ ancestry with LBD in this autopsy series. However, these patterns may reflect recruitment bias rather than true biological variation, as cultural factors may influence research participation and brain donation, highlighting the need for expanded brain bank representation and validation in underrepresented populations.

This study has several limitations inherent to brain bank research. Referral and sampling bias may persist despite the multi-centre design, reflected in a younger age at diagnosis and longer disease duration than reported in population-based cohorts. Pathological staging systems, while invaluable in understanding disease processes, provide relatively coarse measures of disease burden and may vary between pathologists and brain banks. Quantitative approach using whole scanned digital images may improve correlations between pathology and clinical phenotypes. In addition, incomplete documentation of coexisting pathologies, evolving diagnostic frameworks (e.g. ARTAG), variability in staging systems across centers and time periods, and the use of center-specific or broad classifications (e.g. tauopathy) may have influenced prevalence estimates and interpretation.

This study constitutes one of the largest genotyped and genome sequenced movement disorder cohorts integrated with clinical and neuropathological annotation. Our findings highlight the power of large-scale multimodal integration for advancing understanding of movement disorders. This dataset provides a resource for investigating the phenotypic impact of rare variants and variants of uncertain significance, and for identifying novel genotype-phenotype associations. As digital pathology advances, brain banks should adopt more systematic data collection and prioritise inclusion of underrepresented populations to capture disease heterogeneity across ancestries and support pathology-targeted diagnostics and genotype-informed therapies.

## Contributors

LYW drafted the manuscript. LYW and TdT verified and analysed the data reported in this manuscript. HRM conceptualised and supervised the study.

## Declaration of Interest

HRM received grant funding and support for attending meetings or travel from the Michael J Fox Foundation for Parkinson’s Research for this work; received grant funding from the Progressive Supranuclear Palsy Association, Corticobasal Disease Solutions, Drake Foundation, Parkinson’s UK, Medical Research Council UK, and Cure Parkinson’s Trust unrelated to this work; received consulting fees from Roche, Amylyx, and Aprinoia; received honoraria from Kyowa-Kirin, *British Medical Journal*, and the Movement Disorders Society; is a coapplicant on a patent application related to a C9ORF72 method for diagnosing a neurodegenerative disease; and serves on the Cure Progressive Supranuclear Palsy Association Advisory Board, the Association of British Neurologists Movement Disorders Special Interest Group, and the Association of British Neurologists Neurogenetics Advisory Group.

## Data and code availability

Data used in the preparation of this article were obtained from the Global Parkinson’s Genetics Program (GP2; https://gp2.org). Specifically, we used Tier 2 data from GP2 release 10 (https://doi.org/10.5281/zenodo.15748014). Pathological staging and co-pathology data is in Release 11. See eTable 11 for key resources used in this project. All code generated for this article, and the identifiers for all software programs and packages used, are available on GitHub (https://github.com/GP2code/MD-GAP-GP2-CPC) and were given a persistent identifier via Zenodo (10.5281/zenodo.18020958).

## Data Availability

Data used in the preparation of this article were obtained from the Global Parkinson s Genetics Program (GP2; https://gp2.org). Specifically, we used Tier 2 data from GP2 release 10 (https://doi.org/10.5281/zenodo.15748014). Pathological staging and co-pathology data is in Release 11. See eTable 11 for key resources used in this project. All code generated for this article, and the identifiers for all software programs and packages used, are available on GitHub (https://github.com/GP2code/MD-GAP-GP2-CPC) and were given a persistent identifier via Zenodo (10.5281/zenodo.18020958).

## Acknowledgement

We would like to thank all the donors and their families for their invaluable contribution, as well as the collaborating brain banks for their essential support. This research was funded by the UKRI/MRC award - Defining and diagnosing neurodegenerative Movement Disorders through integrated analysis of Genetics And neuroPathology (MD-GAP) (MR/T018569/1), and the Global Parkinson’s Genetic Program (GP2). GP2 is funded by the Aligning Science Across Parkinson’s (ASAP) initiative and implemented by The Michael J. Fox Foundation for Parkinson’s Research (https://gp2.org). For a complete list of GP2 members, see https://gp2.org. This research was funded in part by Aligning Science Across Parkinson’s ASAP-000478 through the Michael J. Fox Foundation for Parkinson’s Research (MJFF). This paper presents independent research supported by the NIHR, Newcastle Biomedical Research Centre (BRC). This research was also supported in part by the Intramural Research Program of the National Institutes of Health (NIH; program #: ZIANS003154). The contributions of the NIH authors were made as part of their official duties as NIH federal employees, are in compliance with agency policy requirements, and are considered words of the United States Government. However, the findings and conclusions presented in this paper are those of the authors and do not necessarily reflect the views of the NIH or the U.S. Department of Health and Human Services. The Arizona Study of Aging and Neurodegenerative Disorders and Brain and Body Donation Program at Banner Sun Health Research Institute has been supported by the National Institute of Neurological Disorders and Stroke (U24 NS072026 National Brain and Tissue Resource for Parkinson’s Disease and Related Disorders), the National Institute on Aging (P30 AG019610 and P30AG072980, Arizona Alzheimer’s Disease Center), the Arizona Department of Health Services (contract 211002, Arizona Alzheimer’s Research Center), the Arizona Biomedical Research Commission (contracts 4001, 0011, 05-901 and 1001 to the Arizona Parkinson’s Disease Consortium) and the Michael J. Fox Foundation for Parkinson’s Research. For the purpose of open access, the author has applied a CC BY public copyright licence to all Author Accepted Manuscripts arising from this submission.

## Notes

### Author Declarations

Ethical approval to coordinate the MD-GAP study was obtained from the University College London Research Ethics Committee (23473/001).

## References

1. Wenning GK, Stankovic I, Vignatelli L, et al. The Movement Disorder Society Criteria for the Diagnosis of Multiple System Atrophy. Mov Disord. 2022;37(6):1131–1148.

2. Höglinger GU, Respondek G, Stamelou M, et al. Clinical diagnosis of progressive supranuclear palsy: The movement disorder society criteria. Mov Disord. 2017;32(6):853–864.

3. Gibb WR, Lees AJ. The relevance of the Lewy body to the pathogenesis of idiopathic Parkinson’s disease. J Neurol Neurosurg Psychiatry. 1988;51(6):745–752.

4. Armstrong MJ, Litvan I, Lang AE, et al. Criteria for the diagnosis of corticobasal degeneration. Neurology. 2013;80(5):496–503.

5. Postuma RB, Berg D, Stern M, et al. MDS clinical diagnostic criteria for Parkinson’s disease. Mov Disord. 2015;30(12):1591–1601.

6. Neumann J, Bras J, Deas E, et al. Glucocerebrosidase mutations in clinical and pathologically proven Parkinson’s disease. Brain. 2009;132(Pt 7):1783–1794.

7. Kalia LV, Lang AE, Hazrati LN, et al. Clinical correlations with Lewy body pathology in LRRK2-related Parkinson disease. JAMA Neurol. 2015;72(1):100–105.

8. Ben-Joseph A, Marshall CR, Lees AJ, Noyce AJ. Ethnic variation in the manifestation of Parkinson’s disease: A narrative review. J Parkinsons Dis. 2020;10(1):31–45.

9. McKeith IG, Boeve BF, Dickson DW, et al. Diagnosis and management of dementia with Lewy bodies. Neurology. 2017;89(1):88–100.

10. Beach TG, Adler CH, Lue L, et al. Unified staging system for Lewy body disorders: correlation with nigrostriatal degeneration, cognitive impairment and motor dysfunction. Acta Neuropathol. 2009;117(6):613–634.

11. Braak H, Del Tredici K, Rüb U, de Vos RAI, Jansen Steur ENH, Braak E. Staging of brain pathology related to sporadic Parkinson’s disease. Neurobiol Aging. 2003;24(2):197–211.

12. Braak H, Alafuzoff I, Arzberger T, Kretzschmar H, Del Tredici K. Staging of Alzheimer disease-associated neurofibrillary pathology using paraffin sections and immunocytochemistry. Acta Neuropathol. 2006;112(4):389–404.

13. Mirra SS, Heyman A, McKeel D, et al. The Consortium to Establish a Registry for Alzheimer’s Disease (CERAD). Part II. Standardization of the neuropathologic assessment of Alzheimer’s disease. Neurology. 1991;41(4):479–486.

14. Thal DR, Rüb U, Orantes M, Braak H. Phases of A beta-deposition in the human brain and its relevance for the development of AD. Neurology. 2002;58(12):1791–1800.

15. The Global Parkinson’s Genetics Program, Leonard HL. Novel Parkinson’s disease genetic risk factors within and across European populations. *medRxiv*. Published online March 17, 2025:2025.03.14.24319455.

16. Towns C, Richer M, Jasaityte S, et al. Defining the causes of sporadic Parkinson’s disease in the global Parkinson’s genetics program (GP2). NPJ Parkinsons Dis. 2023;9(1):131.

17. Towns C, Fang ZH, Tan MMX, et al. Parkinson’s families project: a UK-wide study of early onset and familial Parkinson’s disease. NPJ Parkinsons Dis. 2024;10(1):188.

18. Vitale D, Koretsky MJ, Kuznetsov N, et al. GenoTools: an open-source Python package for efficient genotype data quality control and analysis. G3 (Bethesda). 2025;15(1). doi:10.1093/g3journal/jkae268

19. Parlar SC, Grenn FP, Kim JJ, Baluwendraat C, Gan-Or Z. Classification of GBA1 variants in Parkinson’s disease: The GBA1-PD browser. Mov Disord. 2023;38(3):489–495.

20. Adler CH, Beach TG, Zhang N, et al. Clinical diagnostic accuracy of early/advanced Parkinson disease: An updated clinicopathologic study. Neurol Clin Pract. 2021;11(4):e414–e421.

21. Pottier C, Küçükali F, Baker M, et al. Deciphering distinct genetic risk factors for FTLD-TDP pathological subtypes via whole-genome sequencing. Nat Commun. 2025;16(1):3914.

22. Hughes AJ, Daniel SE, Kilford L, Lees AJ. Accuracy of clinical diagnosis of idiopathic Parkinson’s disease: a clinico-pathological study of 100 cases. J Neurol Neurosurg Psychiatry. 1992;55(3):181–184.

23. Williams-Gray CH, Mason SL, Evans JR, et al. The CamPaIGN study of Parkinson’s disease: 10-year outlook in an incident population-based cohort. J Neurol Neurosurg Psychiatry. 2013;84(11):1258–1264.

24. Fox SH, Luca DG, Postuma RB, et al. Revisiting the 2015 MDS diagnostic criteria for Parkinson disease: insights from autopsy-confirmed cases. NPJ Parkinsons Dis. 2025;11(1):360.

25. Wilkens I, Bebermeier S, Heine J, et al. Multiple system atrophy without dysautonomia: An autopsy-confirmed study. Neurology. 2025;105(11):e214316.

26. Williams DR, Warren JD, Lees AJ. Using the presence of visual hallucinations to differentiate Parkinson’s disease from atypical parkinsonism. J Neurol Neurosurg Psychiatry. 2008;79(6):652–655.

27. Vaughan DP, Fumi R, Theilmann Jensen M, et al. Evaluation of cerebrospinal fluid α-synuclein seed amplification assay in progressive supranuclear palsy and corticobasal syndrome. Mov Disord. 2024;39(12):2285–2291.

28. Simuni T, Chahine LM, Poston K, et al. A biological definition of neuronal α-synuclein disease: towards an integrated staging system for research. Lancet Neurol. 2024;23(2):178–190.

29. Höglinger GU, Adler CH, Berg D, et al. A biological classification of Parkinson’s disease: the SynNeurGe research diagnostic criteria. Lancet Neurol. 2024;23(2):191–204.

30. Hansen D, Ling H, Lashley T, Holton JL, Warner TT. Review: Clinical, neuropathological and genetic features of Lewy body dementias. Neuropathol Appl Neurobiol. 2019;45(7):635–654.

31. Irwin DJ, Grossman M, Weintraub D, et al. Neuropathological and genetic correlates of survival and dementia onset in synucleinopathies: a retrospective analysis. Lancet Neurol. 2017;16(1):55–65.

32. Smith C, Malek N, Grosset K, Cullen B, Gentleman S, Grosset DG. Neuropathology of dementia in patients with Parkinson’s disease: a systematic review of autopsy studies. J Neurol Neurosurg Psychiatry. 2019;90(11):1234–1243.

33. Hipp MS, Kasturi P, Hartl FU. The proteostasis network and its decline in ageing. Nat Rev Mol Cell Biol. 2019;20(7):421–435.

34. Malek N, Weil RS, Bresner C, et al. Features of GBA-associated Parkinson’s disease at presentation in the UK Tracking Parkinson’s study. J Neurol Neurosurg Psychiatry. 2018;89(7):702–709.

35. Tan MMX, Malek N, Lawton MA, et al. Genetic analysis of Mendelian mutations in a large UK population-based Parkinson’s disease study. Brain. 2019;142(9):2828–2844.

36. Adler CH, Beach TG, Shill HA, et al. GBA mutations in Parkinson disease: earlier death but similar neuropathological features. Eur J Neurol. 2017;24(11):1363–1368.

37. Walton RL, Koga S, Beasley AI, et al. Role of GBA variants in Lewy body disease neuropathology. Acta Neuropathol. 2024;147(1):54.

38. Saunders-Pullman R, Mirelman A, Alcalay RN, et al. Progression in the LRRK2-asssociated Parkinson disease population. JAMA Neurol. 2018;75(3):312–319.

39. Rajput A, Dickson DW, Robinson CA, et al. Parkinsonism, Lrrk2 G2019S, and tau neuropathology. Neurology. 2006;67(8):1506–1508.

40. Blauwendraat C, Pletnikova O, Geiger JT, et al. Genetic analysis of neurodegenerative diseases in a pathology cohort. Neurobiol Aging. 2019;76:214.e1–e214.e9.

41. Sekiya H, Franke L, Hashimoto Y, et al. Widespread distribution of α-synuclein oligomers in LRRK2-related Parkinson’s disease. Acta Neuropathol. 2025;149(1):42.

42. Jensen NM, Vitic Z, Antorini MR, Viftrup TB, Parkkinen L, Jensen PH. Abundant non-inclusion α-synuclein pathology in Lewy body-negative LRRK2-mutant cases. Acta Neuropathol. 2025;149(1):41.

