## Supplementary material for "Pathology and genetics in a global cohort of Parkinsonian Disorders": Supp Files

### Table of content

**eMethods** **2**
Study design 2
DNA extraction 3
Genotyping 3
Genome sequencing 3
Quantification of NeuroBooster array probe performance 4
**eTables** **5**
eTable 1. Number of clinically diagnosed movement disorder cases and control by brain bank 5
eTable 2. Correspondence between Lewy Body Pathology staging systems 6
eTable 3. List of genes assessed in short-read Genome Sequencing (n=38) 7
eTable 4. Variants identified from short-read Genome Sequencing (n=17) and additional variants of interest (n=21) analysed in genotyped samples 9
eTable 5. Frequency of primary pathological diagnosis by genetically determined ancestry 11
eTable 6. Distribution of incidental Lewy body cases by Lewy body disease stage 12
eTable 7. Distribution of secondary pathologies by primary pathology groups 12
eTable 8. Distribution of pathology stages by clinical diagnosis in Lewy body diseases 13
eTable 9. Frequency of GBA1 variants aggregated into Gaucher's Disease causing mutations group and Parkinson's Disease risk mutations group by pathological diagnosis 14
eTable 10. Cases with pathogenic LRRK2 mutations 14
eTable 11. Key Resource Table 15
eTable 12. Performance of NeuroBooster Array (NBA) probes compared with short-read Genome Sequencing (GS) for neurodegenerative movement disorder associated variants. 18
eTable 13. Variants with NeuroBooster Array (NBA) probes not detected in short-read Genome Sequencing (GS). 25
eFigure 1. Proportion of LBD and PSP diagnoses across ancestry groups. 26
**References** **27**

#### Pathology and genetics in a global cohort of Parkinsonian Disorders

##### eMethods

##### Study design

Understanding movement disorders through the integrated analysis of Genetics and neuroPathology (MD-GAP) includes 11 brain banks (eTable 1) and is funded by the Medical Research Council UK and Global Parkinson's Genetic Program (GP2, https://gp2.org/). We obtained ethical approval to coordinate the MD-GAP study from the UCL Queen Square Institute of Neurology research ethics committee (23473/001). Each brain bank has local ethics approval for the recruitment, storage and distribution of brain material from brain donors with appropriate consent. We set up data and material sharing agreements with each brain bank in the UK and Australia. An additional contract was signed with the Michael J. Fox Foundation (MJFF), allowing all genetic data generated through this project to be available on Verily Workbench (https://workbench.verily.com/), a cloud-native platform designed to host harmonised data for biomedical research.

We recruited all available subjects from collaborating brain banks with either DNA or frozen tissue for DNA extraction, with clinically or pathologically confirmed diagnoses of movement disorders. Clinical diagnoses include Parkinson's disease (PD), Parkinson's disease dementia (PDD), dementia with Lewy bodies (DLB), progressive supranuclear palsy (PSP), corticobasal syndrome (CBS), multiple system atrophy (MSA), and neurologically healthy controls. Pathological movement disorders diagnoses include Lewy body disorders (LBD), progressive supranuclear palsy, corticobasal degeneration (CBD), multiple system atrophy and Other (Alzheimer's disease, Argyrophilic grain disease, Cerebral amyloid angiopathy, Cerebral Vascular Disease, Chronic Traumatic Encephalopathy, Frontotemporal lobar degeneration, Primary Age Related Tauopathy, Pick's Disease, Tauopathy not otherwise specified, Tumour). Cases with a pathological diagnosis of motor neuron disease or prion disease were excluded from the study.

The summary demographic, clinical, and pathological data were obtained through three sources: direct sharing by the brain banks' manager, extraction from the UK Brain Bank Network (UKBBN) database, and clinical and pathology reports for cases from London Neurodegenerative Diseases Brain Bank (King's College London) and Multiple Sclerosis and Parkinson's Tissue Bank (Imperial College London). Clinical diagnoses were assigned according to these sources, and where available, clinical records were reviewed by MD-GAP's clinical team. Pathological diagnoses were recorded as reported in the pathology reports, preserving the original order of diagnoses. In cases where a formal diagnosis was not provided but semi-quantitative staging was available, a pathological diagnosis was inferred based on the staging data. Some cases had a clinical diagnosis but no corresponding pathological findings; for example, a clinical diagnosis of PD without LBs on Braak staging was recorded as a control. Cases were only recorded as lacking co-pathologies if this was explicitly stated in the reports. To improve transparency and facilitate assessment of inter-center variability, we compiled a table (eTable 12) summarising diagnostic criteria, staging system and antibodies used by the contributing brain banks, where such information was available.

We used REDCap, a secure web-based application designed for data collection and management (https://projectredcap.org/) to store clinical and pathological data. The database includes five instruments: 1) Base and demographics, 2) Family history, 3) Clinical information, 4) Neuropathology and 5) Genetics and transcriptomics. The data fields were harmonised with data fields from the GP2 data dictionary to facilitate data integration and large-scale meta-analyses. GP2 is an ASAP-funded project aimed at genotyping over 150,000 participants and genome sequence (GS) over 10,000 individuals across the world to further our understanding of the genetic architecture of PD. GP2 includes clinical and brain bank cohorts.

##### DNA extraction

DNA was extracted from either the cerebellum or frontal cortex. We conducted quality control by performing Qubit fluorometry or nanodrop (260/280 ratio of ~1.8) to determine DNA concentration and diluted accordingly in 96-well plates before genotyping and sequencing.

##### Genotyping

Available samples were genotyped using the Illumina Global Diversity Array chip with Neurobooster, which has a high-density SNV backbone (1.9 M SNVs) with an extra 95K custom content added, including known causal variants for various neurodegenerative diseases and imputation boosters for underrepresented populations (https://github.com/GP2code/Neuro_Booster_Array). Unlike GS, microarray genotyping focuses on specific regions. Genotyped data were called from IDAT files and processed using GenoTools (https://github.com/GP2code/GenoTools), a pipeline developed to automate the quality control (QC) process and estimation of ancestry. All data were imputed against the TOPMed r2 panel with Eagle v2.4 phasing on the TOPMed Imputation Server using Minimac4 (https://imputation.biodatacatalyst.nhlbi.nih.gov/33).

Genotype data were filtered using PLINK v2 (https://www.cog-genomics.org/plink/2.0/). Individuals and SNPs with >5% missing data were excluded using the –mind 0.05 and –geno 0.05 filters, respectively, to ensure high-quality data.

Variant selection was based on GS findings and additional important movement disorder-associated variants, shown in eTable 4. GS identified variants from this cohort (n=17) classified as pathogenic/ likely pathogenic based on Clinvar (https://www.ncbi.nlm.nih.gov/clinvar/ downloaded on 2024/09/17) were assessed in all cases that underwent NeuroBooster array (NBA). Of the 17 variants detected by GS, 13 had valid probes in the Illumina NBA chip. Additional rare variants (n = 21) of interest in *GBA1*, *LRRK2*, *PINK1*, *PRKN*, *SNCA* and *VPS35* with probes present in the NBA chip were selected. This includes all *LRRK2* and *SNCA* variants defined as pathogenic in Genereviews (https://www.ncbi.nlm.nih.gov/books/NBK1116/) and ClinVar (https://www.ncbi.nlm.nih.gov/clinvar/), *GBA1* variants (PD risk alleles and GD causing variants) that occurred in >0.1% of European PD as per Malek et al. 2018 [1], *PRKN* variants with >1% frequency in autosomal recessive PD patients as per Menon et al. 2024 [2], and the *PINK1* L347P variant described by Morales-Briceno et al. [3]. Therefore, a total of 34 variants were assessed in cases that underwent NBA.

##### Genome sequencing

Selected samples (n = 2954) underwent Illumina short-read Genome sequencing (GS) to examine both coding and non-coding regions, enabling detection of rare and common variants, structural variations, and mutations linked to phenotypes. Reads were aligned to the GRCh38DH reference genome, and small variants were called.

Joint genotyping was performed using the Broad Institute's pipeline for samples passing AMP-PD quality control (https://amp-pd.org). gVCFs generated with DeepVariant were merged using GLnexus (v1.6.1) with settings requiring minimum allele quality scores (min_AQ1, min_AQ2 ≥10), allowing monoallelic sites, and supporting up to 32 alternate alleles. The pipeline preserved partial data, genotype likelihoods (PL), and revised genotypes. Sample-level FORMAT fields—DP, AD, GQ, and PL—were conservatively merged (e.g., minimum DP and GQ values).

High-quality variants were retained if they passed variant quality score recalibration, had a call rate >0.95, genotype quality ≥20 and read depth ≥10. No allele balance (AB) filtering was applied at the sample level. Samples were excluded for excessive heterozygosity (F-statistic > ±0.15) or sex mismatches between clinical and genetic sex (based on the X chromosome). Relatedness checks were used to identify duplicates and contamination.

A total of 415 genes were selected based on the Genomics England PanelApp Adult Onset Neurodegenerative Disorders panel, release R.58 (v7.15, 30 October 2024) https://panelapp.genomicsengland.co.uk/panels/474/. Gene-level variant extraction was performed using bcftools (bcftools(1)) and variant-level annotation was performed with ANNOVAR (December 2023, ANNOVAR Documentation). Only genes associated with a movement disorder phenotype and a "green" (GEL status 3) classification, indicating a high level of evidence supporting the gene's role in disease, were included in the final analysis (n=36). Exceptions were made for genes of particular interest regardless of GEL status, including RAB39B and VPS13C, therefore, 38 genes were examined as described in eTable 3.

Variants in dominant and recessive genes were retained if coding or splicing, and classified as pathogenic/ likely pathogenic according to Clinvar (https://www.ncbi.nlm.nih.gov/clinvar/). *GBA1* variants were retained irrespective of Clinvar classification and allele frequency. Single heterozygous carriers in recessive genes were excluded from analysis, except for *GBA1*. Synonymous variants were excluded.

17 variants of interest were identified in 2954 cases, shown in eTable 4.

**Quantification of NeuroBooster array probe performance**

In a subset of cases (n=2214) across varied cohorts both NBA and GS data were available. We calculated the proportion of false positive and false negative genotype calls and calculated the sensitivity and specificity of each probe for variants detected by NBA, considering GS as the reference (eTable 13). When more than one probe was available for the same variant, the best probe was selected, prioritising specificity over sensitivity to limit the rate of false positives. Where probes performed equally, a single probe per variant was selected. Of the variants searched for in NBA, 21 probes were validated against GS, each demonstrating a specificity greater than 99.9%.

eTable 14 lists NBA probe names for variants not detected in GS. As these variants were not observed in GS, their corresponding probes were not validated and were not reported further.

Combining GS and validated findings genotyping, 20 unique mutations were identified in our cohort. Following collaboration with brain banks and review of relevant literature, variants in *PSAP* and *C19orf12* were excluded from the main results shown in Table 3. In contrast to the classification in PanelApp, we found no confirmatory segregation studies for *PSAP*, and reported cases have largely involved sporadic rather than familial disease [4]. Furthermore, neither of the two cases with *C19orf12* variants demonstrated brain iron accumulation on post-mortem examination, questioning its pathogenicity. Accordingly, only 18 pathogenic or likely pathogenic variants are reported in the main text.

#### eTables

##### eTable 1. Number of clinically diagnosed movement disorder cases and control by brain bank

| Brain Bank | Contacts | N |
|---|---|---|
| Queen Square Brain Bank | **Zane Jaunmuktane**<br>**Tammaryn Lashley**<br>**Tom Warner** | 1335 |
| Multiple Sclerosis and Parkinson's Tissue Bank | Djordje Gveric<br>**Steve Gentleman** | 637 |
| Banner Sun Health Research Institute [5] | **Thomas Beach**<br>Geidy Serrano | 551 |
| Edinburgh Brain and Tissue Bank | **Colin Smith**<br>Chris-Anne McKenzi | 138 |
| Newcastle Brain Tissue Resource | Debbie Lett<br>**Chris Morris** | 113 |
| Oxford Brain Bank | Carolyn Sloan<br>**Laura Parkkinen** | 116 |
| Sydney Brain Bank | **Glenda Halliday**<br>**Claire Shepherd** | 110 |
| Manchester Brain Bank | **Andrew Robinson**<br>**Federico Roncaroli** | 110 |
| London Neurodegenerative Diseases Brain Bank | Claire Troakes<br>**Andrew King** | 101 |
| South West Dementia Brain Bank | Candida Tasman<br>Richard Cain<br>**Seth Love** | 78 |
| Victoria Brain Bank | **Catriona McLean** | 64 |
| **Total** | - | 3353 |

11 brain banks have been included in the present study. UK and Australian brain banks are part of the MD-GAP study. Banner Sun Health Research Institute is part of the GP2 study only.
In the Contacts column, group Principal Investigators are in bold

#### eTable 2. Correspondence between Lewy Body Pathology staging systems

| Unified staging system for LBD | McKeith stage | Lewy Body Braak stage |
|---|---|---|
| Neocortical | Neocortical | Stages 5 - 6 |
| Limbic | Limbic | Stage 4 |
| Brainstem | Brainstem | Stages 1 - 3 |
| Amygdala | Amygdala | .. |

A table showing equivalence between different staging systems. Based on BrainNet Europe Protocol [6].

**eTable 3. List of genes assessed in short-read Genome Sequencing (n=38)**

| Gene | Mode of Inheritance | Phenotypes |
|---|---|---|
| *ATP1A3* | Monoallelic | Alternating hemiplegia of childhood; CAPOS syndrome; Rapid-Onset Dystonia-Parkinsonism |
| *AUH* | Biallelic | Dystonia |
| *C19orf12* | Both Monoallelic and Biallelic | Spastic paraplegia; Neurodegeneration with brain iron accumulation |
| *CHCHD2* | Monoallelic | PD |
| *CHMP2B* | Monoallelic | FTD and/or amyotrophic lateral sclerosis; Dystonia |
| *COASY* | Biallelic | COASY protein-associated neurodegeneration; Neurodegeneration with brain iron accumulation |
| *DNAJC6* | Biallelic | EOPD |
| *FBXO7* | Biallelic | Dystonia; PD |
| *FTL* | Monoallelic | Neurodegeneration with brain iron accumulation |
| *GBA* | Biallelic | Late-onset PD |
| *GCH1* | Both Monoallelic and Biallelic | Dystonia; DOPA-responsive; with or without hyperphenylalaninemia |
| *KIAA1161* | Biallelic | Basal ganglia calcification |
| *LRRK2* | Monoallelic | PD |
| *LYST* | Biallelic | Chediak-Higashi syndrome; peripheral neuropathy; Parkinsonism; spastic paraplegia |
| *MAPT* | Monoallelic | FTD with or without parkinsonism |
| *NAA60* | Biallelic | Basal ganglia calcification |
| *NPC1* | Biallelic | Niemann-Pick disease |
| *NPC2* | Biallelic | Dystonia; Niemann-Pick disease |
| *PANK2* | Biallelic | Dystonia; Neurodegeneration with brain iron accumulation |

| Gene | Mode of Inheritance | Phenotypes |
|---|---|---|
| ***PARK7*** | Biallelic | EOPD |
| ***PDGFB*** | Monoallelic | Basal ganglia calcification |
| ***PDGFRB*** | Monoallelic | Dystonia; Basal ganglia calcification |
| ***PINK1*** | Biallelic | EOPD; Dystonia |
| **Gene** | **Mode of Inheritance** | **Phenotypes** |
| ***PLA2G6*** | Biallelic | PD; Neurodegeneration with brain iron accumulation |
| ***PRKN*** | Biallelic | PD |
| ***PSAP*** | Monoallelic | PD |
| ***RAB32*** | Monoallelic | PD |
| ***RAB39B*** | X-Linked: hemizygous mutation in males, monoallelic mutations in females may cause disease (may be less severe, later onset than males) | Early onset parkinsonism and intellectual disability; Waisman syndrome |
| ***SLC20A2*** | Monoallelic | Dystonia; Basal ganglia calcification |
| ***SNCA*** | Monoallelic | PD; Dementia |
| ***SPG11*** | Biallelic | Early onset parkinsonism; Spastic paraplegia |
| ***SYNJ1*** | Biallelic | PD |
| ***TBK1*** | Monoallelic | FTD and/or ALS; PSP-like and Cerebellar phenotypes |
| ***TREM2*** | Biallelic | Polycystic lipomembranous osteodysplasia with sclerosing leukoencephalopathy; Dystonia |
| ***VPS13C*** | Biallelic | EOPD |
| ***VPS35*** | Monoallelic | PD |
| ***WDR45*** | X-Linked: hemizygous mutation in males | Dystonia;Neurodegeneration with brain iron accumulation |
| ***XPR1*** | Monoallelic | Basal ganglia calcification |

Abbreviations: PD: Parkinson disease; EOPD: Early onset Parkinson disease; LOPD: Late onset Parkinson disease; FTD: Frontotemporal dementia; ALS: Amyotrophic lateral sclerosis; PSP: Progressive Supranuclear Palsy

**eTable 4. Variants identified from short-read Genome Sequencing (n=17) and additional variants of interest (n=21) analysed in genotyped samples**

| Gene | Hg38 Chr:pos:ref:alt | HGVS | NBA Illumina probe present (Y/N) | Variant selection |
|---|---|---|---|---|
| *C19orf12* | 19:29702957:29702967:ACAGCCCCCC G:- | p.Gly58ArgfsTer10 | N | Short-read GS |
| *GBA1* | 1:155235843:T:C | p.Asn409Ser | Y | Short-read GS |
| *GBA1* | 1:155236246:G:A | p.Thr408Met | Y | Short-read GS |
| *GBA1* | 1:155236376:C:T | p.Glu365Lys | Y | Short-read GS |
| *GBA1* | 1:155238192:A:G | p.Ser235Pro | Y | Short-read GS |
| *GBA1* | 1:155237444:A:G | p.Ile299Thr | Y | Short-read GS |
| *GBA1* | 1:155235252:A:G | p.Leu483Pro | Y | Short-read GS |
| *GBA1* | 1:155237426:G:- | p.Pro305LeufsTer 31 | N | Short-read GS |
| *GBA1* | 1:155238630:G:A | p.Arg159Trp | N | Short-read GS |
| *GBA1* | 1:155235727:C:G | p.Asp448His | Y | Short-read GS |
| *GBA1* | 1:155240629:C:T | .. | Y | Short-read GS |
| *GBA1* | 1:155236384:G:A | p.Thr362Ile | Y | Short-read GS |
| *LRRK2* | 12:40340400:G:A | p.Gly2019Ser | Y | Short-read GS |
| *LRRK2* | 12:40321114:A:G | p.Tyr1699Cys | Y | Short-read GS |
| *PRKN* | 6:161785820:G:A | p.Arg275Trp | Y | Short-read GS |
| *PSAP* | 10:71851221:T:A | p.Met1Leu | Y | Short-read GS |
| *TBK1* | 12:64498008:G:T | p.Glu703Ter | N | Short-read GS |
| *GBA1* | 1:155235196:G:A | p.Arg502Cys | Y | Additional variant of interest |

| Gene | Hg38 Chr:pos:ref:alt | HGVS | NBA Illumina probe present (Y/N) | Variant selection |
|---|---|---|---|---|
| *GBA1* | 1:155238597:G:A | p.Arg170Cys | Y | Additional variant of interest |
| *PINK1* | 1:20645640:T:C | p.Leu347Pro | Y | Additional variant of interest |
| *PRKN* | 1:34784887:G:A | p.Arg42Pro | Y | Additional variant of interest |
| *LRRK2* | 12:40299125:A:G | p.Ile1122Val | Y | Additional variant of interest |
| *LRRK2* | 12:40309225:A:G | p.Asn1437Asp | Y | Additional variant of interest |
| *LRRK2* | 12:40310434:C:T | p.Arg1441Cys | Y | Additional variant of interest |
| *LRRK2* | 12:40310435:G:A | p.Arg1441His | Y | Additional variant of interest |
| *LRRK2* | 12:40340404:T:C | p.Ile2020Thr | Y | Additional variant of interest |
| *LRRK2* | 12:40363526:G:A | p.Gly2385Asp | Y | Additional variant of interest |
| *VPS35* | 16:46662452:C:T | p.Asp620Asn | Y | Additional variant of interest |
| *SNCA* | 4:89828149:C:T | p.Ala53Thr | Y | Additional variant of interest |
| *SNCA* | 4:89828154:C:T | p.Gly51Asp | Y | Additional variant of interest |
| *SNCA* | 4:89828170:C:T | p.Glu46Lys | Y | Additional variant of interest |
| *SNCA* | 4:89835580:C:G | p.Ala30Pro | Y | Additional variant of interest |
| *PRKN* | 6:161350205:C:T | p.Cys431Phe | Y | Additional variant of interest |
| *PRKN* | 6:161350208:C:T | p.Gly430Asp | Y | Additional variant of |

| Gene | Hg38 Chr:pos:ref:alt | HGVS | NBA Illumina probe present (Y/N) | Variant selection |
|---|---|---|---|---|
| | | | | interest |
| *PRKN* | 6:161360190:G:T | p.Glu395Ter | Y | Additional variant of interest |
| *PRKN* | 6:161785885:C:T | p.Cys253Tyr | Y | Additional variant of interest |
| *PRKN* | 6:162262715:C:CCA | p.Trp74CysfsTer8 | Y | Additional variant of interest |
| *PRKN* | 6:162443378:CCT:C | p.Gln34ArgfsTer5 | Y | Additional variant of interest |

eTable 4 contains all neurodegenerative movement disorder associated variants analysed in genotyped samples. Variants were selected from two sources: (1) ClinVar pathogenic/likely pathogenic variants identified from short-read genome sequencing (GS) (n=17; 13 with valid Illumina NBA chip probes), and (2) additional rare variants of interest (n=21) in *GBA1, LRRK2, PINK1, PRKN, SNCA,* and *VPS35.* A total of 34 variants were evaluated in NBA-genotyped cases.

#### eTable 5. Frequency of primary pathological diagnosis by genetically determined ancestry

| | AAC/AFR | AJ | AMR | CAH | CAS | EAS | EUR | MDE | SAS |
|---|---|---|---|---|---|---|---|---|---|
| **N*** | 5 | 63 | 8 | 9 | 1 | 3 | 2758 | 2 | 18 |
| **Sex, F (%)** | 1 (20.0%) | 25 (39.7%) | 3 (37.5%) | 3 (33.3%) | 0 (0.0%) | 2 (66.7%) | 1051 (38.1%) | 1 (50.0%) | 8 (44.4%) |
| **Age at death, years(sd)** | 71.2 (3.7) | 79.9 (9.1) | 73.8 (12.7) | 73.2 (9.0) | 53 | 72.7 (1.5) | 77.2 (10.2) | 81.5 (7.8) | 71.1 (4.9) |
| **Pathology** | | | | | | | | | |
| LBD | 3 (60.0%) | 40 (63.5%) | 1 (12.5%) | 1 (11.1%) | 0 (0.0%) | 1 (33.3%) | 1510 (54.7%) | 1 (50.0%) | 2 (11.1%) |
| PSP | 2 (40.0%) | 5 (7.9%) | 1 (12.5%) | 5 (55.6%) | 0 (0.0%) | 1 (33.3%) | 433 (15.7%) | 1 (50.0%) | 12 (66.7%) |
| CBD | 0 (0.0%) | 0 (0.0%) | 1 (12.5%) | 0 (0.0%) | 0 (0.0%) | 1 (33.3%) | 14 (0.5%) | 0 (0.0%) | 0 (0.0%) |
| MSA | 0 (0.0%) | 3 (4.8%) | 0 (0.0%) | 0 (0.0%) | 1 (100.0%) | 0 (0.0%) | 187 (6.8%) | 0 (0.0%) | 2 (11.1%) |

| | | | | | | | | | |
|---|---|---|---|---|---|---|---|---|---|
| Control | 0 (0.0%) | 13 (20.6%) | 4 (50.0%) | 2 (22.2%) | 0 (0.0%) | 0 (0.0%) | 518 (18.8%) | 0 (0.0%) | 1 (5.5%) |
| Other | 0 (0.0%) | 2 (3.2%) | 1 (12.5%) | 1 (11.1%) | 0 (0.0%) | 0 (0.0%) | 96 (3.5%) | 0 (0.0%) | 1 (5.5%) |

AAC:African American, AFR:African, AJ:Ashkenazi Jews, AMR:Admixed American/Latin American, CAH:Complex Admixture History, CAS:Central Asian, EAS:East Asian, EUR: European, MDE: Middle Eastern, SAS: South Asian
LBD: Lewy Body Diseases,PSP: Progressive Supranuclear Palsy, MSA: Multiple System Atrophy, CBD: Corticobasal Degeneration

#### eTable 6. Distribution of incidental Lewy body cases by Lewy body disease stage

| LBD Subtype | N | N with *APOE* e4 |
|---|---|---|
| Amygdala | 1 (3.0%) | 0 (0.0%) |
| Brainstem | 7 (21.2%) | 0 (0.0%) |
| Limbic | 7 (21.2%) | 0 (0.0%) |
| Neocortical | 10 (30.3%) | 2 (40.0%) |
| No information available | 5 (15.2%) | 1 (20.0%) |
| *Unclassifiable | 3 (9.1%) | 2 (40.0%) |
| Total | 33 | 5 (15.2%) |

*Cases listed as “Unclassifiable” were designated as such according to the Braak LB system

#### eTable 7. Distribution of secondary pathologies by primary pathology groups

| Secondary Pathology | | Primary Pathology | | | |
|---|---|---|---|---|---|
| | | LBD | PSP | MSA | CBD |
| | n* | 1064 | 216 | 20 | 12 |
| | None | 209 (19.6%) | 1 (0.5%) | 0 (0.0%) | 0 (0.0%) |

| | | | | | |
|---|---|---|---|---|---|
| | **LBD** | .. | 40 (18.5%) | 4 (20.0%) | 3 (25.0%) |
| | **AD** | 426 (40.0%) | 35 (16.2%) | 4 (20.0%) | 2 (16.7%) |
| | **PSP** | 13 (1.2%) | .. | 0 (0.0%) | 0 (0.0%) |
| | **MSA** | 5 (0.5%) | 0 (0.0%) | .. | 0 (0.0%) |
| | **CBD** | 3 (0.3%) | 4 (1.9%) | 0 (0.0%) | .. |
| | **Other** | 408 (38.3%) | 136 (63.0%) | 12 (60.0%) | 7 (58.3%) |

* This table presents only cases for which data on secondary pathology were available.
Other tauopathies: Argyrophilic grain disease, Chronic Traumatic Encephalopathy, Primary Age Related Tauopathy, Picks, Tauopathy, Aging-related tau astrogliopathy
Other: Cerebral amyloid angiopathy, Other, Limbic-predominant age-related TDP-43 encephalopathy, Frontotemporal Lobar Degeneration, tumour

#### eTable 8. Distribution of pathology stages by clinical diagnosis in Lewy body diseases

| | | PD | PDD | DLB |
|---|---|---|---|---|
| McKeith Stages | **Total** | 744 | 288 | 140 |
| | **None** | 14 (1.9%) | 2 (0.7%) | 3 (2.1%) |
| | **Amygdala** | 6 (0.8%) | 3 (1.0%) | 0 (0.0%) |
| | **Brainstem** | 128 (17.2%) | 48 (16.7%) | 14 (10.0%) |
| | **Limbic** | 200 (26.9%) | 41 (14.2%) | 10 (7.1%) |
| | **Neocortical** | 396 (53.2%) | 194 (67.4%) | 113 (80.7%) |
| Braak LB | **Total** | 582 | 237 | 80 |

| | | | | |
|---|---|---|---|---|
| | **0** | 32 (5.4%) | 7 (3.0%) | 2 (2.5%) |
| | **1-2** | 14 (2.5%) | 3 (1.2%) | 0 (0.0%) |
| | **3-4** | 149 (25.6%) | 27 (11.4%) | 2 (2.5%) |
| | **5-6** | 387 (66.5%) | 200 (84.4%) | 76 (95.0%) |
| | **Total** | 778 | 351 | 178 |
| | **0** | 55 (7.1%) | 18 (5.1%) | 5 (2.8%) |
| | **I-II** | 404 (51.9%) | 187 (53.3%) | 63 (35.4%) |
| | **III-IV** | 258 (33.2%) | 118 (33.6%) | 82 (46.1%) |
| **Braak NFT** | **V-VI** | 61 (7.8%) | 28 (8.0%) | 28 (15.7%) |
| | **Total** | 703 | 258 | 123 |
| | **No neuritic plaques** | 388 (55.2%) | 133 (51.6%) | 25 (20.3%) |
| | **Sparse neuritic plaques** | 119 (16.9%) | 64 (24.8%) | 16 (13.0%) |
| | **Moderate neuritic plaques** | 113 (16.1%) | 46 (17.8%) | 28 (22.8%) |
| **CERAD** | **Frequent neuritic plaques** | 83 (11.8%) | 15 (5.8%) | 54 (43.9%) |
| | **Total** | 615 | 248 | 121 |
| | **0** | 159 (26.0%) | 77 (31.1%) | 17 (14.0%) |
| | **1-2** | 197 (32.0%) | 69 (27.8%) | 21 (17.4%) |
| **Thal** | **3** | 151 (24.5%) | 55 (22.2%) | 50 (41.3%) |

| | | | | |
|---|---|---|---|---|
| | 4-5 | 108 (17.5%) | 47 (18.9%) | 33 (27.3%) |

Percentages are calculated using cases with available data for each staging system.
PD: Parkinson's Disease, PDD: Parkinson's Disease Dementia, DLB: Dementia with Lewy Bodies

#### eTable 9. Frequency of *GBA1* variants aggregated into Gaucher's Disease causing mutations group and Parkinson's Disease risk mutations group by pathological diagnosis

| | LBD | PSP | CBD | MSA | Other | All Cases | Control | TOTAL |
|---|---|---|---|---|---|---|---|---|
| WGS N | 1157 | 473 | 20 | 183 | 76 | 1909 | 387 | 2296 |
| *GBA1* GD variants | 33 (2.9%) | 1 (0.2%) | 0 | 1 (0.5%) | 0 | 35 (1.8%) | 2 (0.5%) | 37 (1.6%) |
| *GBA1* Risk variants | 131 (11.3%) | 27 (5.7%) | 1 (5.0%) | 9 (4.9%) | 5 (6.6%) | 173 (9.1%) | 33 (8.5%) | 206 (9.0%) |

LBD: Lewy Body Diseases,PSP: Progressive Supranuclear Palsy, MSA: Multiple System Atrophy, CBD: Corticobasal Degeneration
Other: Alzheimer's disease, Argyrophilic grain disease, Cerebral amyloid angiopathy, Cerebral Vascular Disease, Chronic Traumatic Encephalopathy, Frontaltemporal lobar degeneration, Primary Age Related Tauopathy, Pick's Disease, Small Vessels Disease, Tauopathy, Tumour
*GBA1* GD causing mutations include: D448H, I299T, L483P, N409S, P305Lfs*31, R159W, R170C, R502C, S235P, T362I. GBA1 PD risk mutations include: E365K and T408M.

#### eTable 10. Cases with pathogenic *LRRK2* mutations

| Case | Sex | Genetic ancestry | Clinical Diagnosis | Pathological Diagnosis | Age at death | Mutation |
|---|---|---|---|---|---|---|
| 1 | M | EUR | PD | LBD | 81 - 85 | G2019S |
| 2 | F | AJ | PD | LBD | 81 - 85 | G2019S |
| 3 | F | EUR | PD | PSP | 91 - 95 | G2019S |
| 4 | F | EUR | PD | Control | 76 - 80 | G2019S |
| 5 | F | EUR | PD | LBD | 71 - 75 | G2019S |
| 6 | M | EUR | PD | LBD | 76 - 80 | G2019S |

| | | | | | | |
|---|---|---|---|---|---|---|
| 7 | F | AJ | PD | LBD | 81 - 85 | G2019S |
| 8 | M | EUR | PSP | FTLD | 71 - 75 | G2019S |
| 9 | M | CAH | PD | LBD | 55 – 60 | G2019S |
| 10 | F | EUR | PD | PSP | 81 - 85 | G2019S |
| 11 | F | EUR | PD | LBD | 81 - 85 | G2019S |
| 12 | F | EUR | PDD | TDP-43 | 86 - 90 | G2019S |
| 13 | F | AJ | PD | LBD | 81 - 85 | G2019S |
| 14 | F | EUR | PD | Other | 71 - 75 | Y1699C |
| 15 | M | AJ | PD | LBD | 61 - 65 | G2019S |
| 16 | F | EUR | PD | LBD | 81 - 85 | G2019S |
| 17 | F | EUR | PD | LBD | 71 - 75 | G2019S |
| 18 | M | EUR | PD | LBD | 71 - 75 | G2019S |
| 19 | F | EUR | PD | LBD | 86 - 90 | G2019S |

#### eTable 11. Key Resource Table

| RESOURCE TYPE | RESOURCE NAME | SOURCE | IDENTIFIER | NEW/REUSE | ADDITIONAL INFORMATION |
|---|---|---|---|---|---|
| Dataset | Defining and Diagnosing neurodegenerative Movement Disorders through integrated analysis of Genetics and neuroPathology (MD-GAP)/Global Parkinson's | AMP-PD | DOI: 10.1002/mds.28494 | NEW | All data generated in MD-GAP are now part of GP2. Full data can be accessed here: https://amp-pd.org/register-for-amp-pd |

| | | | | |
|---|---|---|---|---|
| | Genetics Program (GP2) | | | |
| Software/code | R code: Analysis Script | Github | https://github.com/huw-morris-lab/MD-GAP-GP2-CPC.git | NEW |
| Software/code | GenoTools | https://github.com/dvitale199/GenoTools | https://doi.org/10.1093/g3journal/jkae268 | REUSE |
| Software/code | R Project for Statistical Computing, v4.3.1 | https://www.r-project.org/ | RRID:SCR_001905 | REUSE |
| Software/code | R package: dplyr | https://cran.r-project.org/web/packages/dplyr/index.html | RRID:SCR_016708 | REUSE |
| Software/code | R package: data.table | https://github.com/Rdatatable/data.table | RRID:SCR_026117 | REUSE |
| Software/code | R package: readxl | https://cran.r-project.org/web/packages/readxl/index.html | RRID:SCR_018083 | REUSE |
| Software/code | R package: stringr | https://stringr.tidyverse.org/ | RRID:SCR_022813 | REUSE |
| Software/code | R package: ggplot2 | https://cran.r-project.org/web/packages/ggplot2/index.html | RRID:SCR_014601 | REUSE |
| Software/code | R package: scales | https://CRAN.R-project.org/package=scales | RRID:SCR_019295 | REUSE |
| Software/code | R package: RColorBrewer | https://cran.r-project.org/web/packages/RColorBrewer/index.html | RRID:SCR_016697 | REUSE |
| Software/code | R package: tidyverse | https://CRAN.R- | RRID:SCR_019186 | REUSE |

| | | | | |
|---|---|---|---|---|
| | | project.org/package=tidyverse | | |
| Software/code | R package: pheatmap | https://www.rdocumentation.org/packages/pheatmap/versions/0.2/topics/pheatmap | RRID:SCR_016418 | REUSE |
| Software/code | R package: gridExtra | https://CRAN.R-project.org/package=gridExtra | RRID:SCR_025249 | REUSE |
| Software/code | R package: MASS | https://CRAN.R-project.org/package=MASS | RRID:SCR_019125 | REUSE |
| Software/code | R package: survival | https://CRAN.R-project.org/view=Survival | RRID:SCR_026244 | REUSE |
| Software/code | R package: survminer | https://rdocumentation.org/packages/survminer/versions/0.4.9 | RRID:SCR_021094 | REUSE |
| Software/code | R package: broom | https://cran.r-project.org/web/packages/broom/index.html | RRID:SCR_026874 | REUSE |
| Software/code | Plink2 | https://www.cog-genomics.org/plink/2.0/ | RRID:SCR_001757 | REUSE |

List of resources generated and used for this project.

**eTable 12. Performance of NeuroBooster Array (NBA) probes compared with short-read Genome Sequencing (GS) for neurodegenerative movement disorder associated variants.**

| Gene | HGVS | Illumina probe name | TP | TN | FP | FN | Sensitivity | Specificity |
|---|---|---|---|---|---|---|---|---|
| *GBA1* | p.Asp448His | **1:155205518-G-C** | 0 | 2211 | 0 | 3 | 0 | 1 |
| *GBA1* | p.Asp448His | ilmnseq_rs1064651 | 0 | 2211 | 0 | 3 | 0 | 1 |
| *GBA1* | p.Asp448His | ilmnseq_rs1064651_ilmndup2_ilmnbot | 0 | 2211 | 0 | 3 | 0 | 1 |
| *GBA1* | p.Asp448His | rs1064651 | 0 | 2211 | 0 | 3 | 0 | 1 |
| *GBA1* | p.Glu365Lys | **chr1:155236376:C:T** | 93 | 2119 | 1 | 1 | 0.989361702 | 0.999528302 |
| *GBA1* | p.Glu365Lys | kgp308887 | 93 | 2119 | 1 | 1 | 0.989361702 | 0.999528302 |
| *GBA1* | p.Ile299Thr | seq-rs794727908 | 0 | 2213 | 1 | 0 | - | 0.999548329 |
| *GBA1* | p.Ile299Thr | **Seq_rs794727908** | 0 | 2214 | 0 | 0 | - | 1 |

| | | | | | | | | |
|---|---|---|---|---|---|---|---|---|
| *GBA1* | p.Asn409Ser | exm106217 | 12 | 2199 | 1 | 2 | 0.8571 42857 | 0.9995 45455 |
| *GBA1* | p.Asn409Ser | ilmnseq_rs76763715.1_F2BT | 9 | 2199 | 1 | 5 | 0.6428 57143 | 0.9995 45455 |
| *GBA1* | p.Asn409Ser | ilmnseq_rs76763715.2_F2BT | 0 | 2200 | 0 | 14 | 0 | 1 |
| *GBA1* | p.Asn409Ser | Seq_rs76763715.2_ilmnfwd_ilmnF2BT | 0 | 2200 | 0 | 14 | 0 | 1 |
| *GBA1* | p.Asn409Ser | **chr1:155235843:T:C** | 14 | 2198 | 2 | 0 | 1 | 0.9990 90909 |
| *GBA1* | p.Arg170Cys | **chr1:155208388-155208388_G_T** | 0 | 2212 | 0 | 2 | 0 | 1 |
| *GBA1* | p.Arg170Cys | rs398123530 | 0 | 2212 | 0 | 2 | 0 | 1 |
| *GBA1* | p.Arg502Cys | **1:155204987-C-T** | 4 | 2210 | 0 | 0 | 1 | 1 |
| *GBA1* | p.Arg502Cys | Seq_rs80356771.1_ilmnrev_ilmnF2BT | 4 | 2210 | 0 | 0 | 1 | 1 |

| | | | | | | | | |
|---|---|---|---|---|---|---|---|---|
| *GBA1* | p.Arg502Cys | Seq_rs80356771.2_ilmnrev_ilmnF2BT | 0 | 2210 | 0 | 4 | 0 | 1 |
| *GBA1* | p.Arg502Cys | rs80356771 | 4 | 2210 | 0 | 0 | 1 | 1 |
| *GBA1* | c.115+1G>A | **rs104886460** | 0 | 2213 | 0 | 1 | 0 | 1 |
| *GBA1* | p.Thr362Ile | **1:155206175-C-T** | 1 | 2213 | 0 | 0 | 1 | 1 |
| *GBA1* | p.Thr362Ile | rs76539814 | 0 | 2213 | 0 | 1 | 0 | 1 |
| *GBA1* | p.Thr362Ile | Seq_rs76539814 | 1 | 2213 | 0 | 0 | 1 | 1 |
| *GBA1* | p.Thr408Met | exm106220 | 42 | 2167 | 2 | 3 | 0.933333333 | 0.999077916 |
| *GBA1* | p.Thr408Met | rs75548401 | 39 | 2167 | 2 | 6 | 0.866666667 | 0.999077916 |
| *GBA1* | p.Thr408Met | **seq_rs75548401** | 43 | 2167 | 2 | 2 | 0.955555556 | 0.999077916 |
| *LRRK2* | p.Gly2019Ser | **exm994671** | 12 | 2202 | 0 | 0 | 1 | 1 |

| | | | | | | | | |
|---|---|---|---|---|---|---|---|---|
| *LRRK2* | p.Gly2019Ser | rs34637584 | 12 | 2202 | 0 | 0 | 1 | 1 |
| *LRRK2* | p.Gly2019Ser | seq_rs34637584 | 12 | 2202 | 0 | 0 | 1 | 1 |
| *LRRK2* | p.Gly2385Asp | **exm994721** | 0 | 2214 | 0 | 0 | - | 1 |
| *LRRK2* | p.Gly2385Asp | rs34778348 | 0 | 2214 | 0 | 0 | - | 1 |
| *LRRK2* | p.Gly2385Asp | seq_rs34778348 | 0 | 2214 | 0 | 0 | - | 1 |
| *LRRK2* | p.Arg1441His | **12:40704237-G-A** | 0 | 2214 | 0 | 0 | - | 1 |
| *LRRK2* | p.Arg1441His | Seq_rs34995376 | 0 | 2214 | 0 | 0 | - | 1 |
| *LRRK2* | p.Tyr1699Cys | **12:40714916-A-G** | 1 | 2213 | 0 | 0 | 1 | 1 |
| *LRRK2* | p.Tyr1699Cys | Seq_rs35801418 | 1 | 2213 | 0 | 0 | 1 | 1 |

| | | | | | | | | |
|---|---|---|---|---|---|---|---|---|
| *PRKN* | p.Cys253Tyr | **PARK2:NM_004562.2:c.758 G>T:p.(Cys253Phe)** | 0 | 2214 | 0 | 0 | - | 1 |
| *PRKN* | p.Gly430Asp | **Seq_rs191486604** | 2 | 2212 | 0 | 0 | 1 | 1 |
| *PRKN* | p.Gly430Asp | exm593474 | 2 | 2212 | 0 | 0 | 1 | 1 |
| *PRKN* | p.Asn52Metf sTer24 | **Variant13588** | 1 | 2213 | 0 | 0 | 1 | 1 |
| *PRKN* | p.Asn52Metf sTer24 | Seq_rs754809877 | 1 | 2213 | 0 | 0 | 1 | 1 |
| *PRKN* | p.Arg275Trp | exm593505 | 19 | 2194 | 0 | 1 | 0.95 | 1 |
| *PRKN* | p.Arg275Trp | **rs34424986** | 20 | 2194 | 0 | 0 | 1 | 1 |
| *PRKN* | p.Arg275Trp | Seq_rs34424986.1_ilmnrev_il mnF2BT | 6 | 2194 | 0 | 14 | 0.3 | 1 |
| *PRKN* | p.Arg275Trp | Seq_rs34424986.2_ilmnrev_il mnF2BT | 0 | 2194 | 0 | 20 | 0 | 1 |
| *PRKN* | p.Trp74Cysfs | **indel.94051** | 1 | 2213 | 0 | 0 | 1 | 1 |

| | | | | | | | | |
|---|---|---|---|---|---|---|---|---|
| | Ter8 | | | | | | | |
| *PSAP* | p.Met1Leu | **10:73610978-A-T** | 1 | 2213 | 0 | 0 | 1 | 1 |
| *PSAP* | p.Met1Leu | Seq_rs121918106.1_ilmnrev_ ilmnF2BT | 1 | 2213 | 0 | 0 | 1 | 1 |
| *PSAP* | p.Met1Leu | Seq_rs121918106.2_ilmnrev_ ilmnF2BT | 0 | 2212 | 1 | 1 | 0 | 0.9995 48125 |
| *SNCA* | p.Gly51Asp | **seq-rs431905511-B1** | 1 | 2213 | 0 | 0 | 1 | 1 |
| *VPS35* | p.Asp620Asn | **16:46696364-G-A** | 0 | 2214 | 0 | 0 | - | 1 |

The proportion of true positive (TP), true negative (TN), false positive (FP) and false negative (FN) genotype calls, and probe sensitivity and specificity, were calculated for the 51 probes targeting mutations detected in the 2214 individuals who underwent both NeuroBooser Array (NBA) genotyping and short-read Genome Sequencing. The most accurate probes were selected for analysis, shown in bold.

**eTable 13. Variants with NeuroBooster Array (NBA) probes not detected in short-read Genome Sequencing (GS).**

| Gene | HGVS | Illumina probe name |
|---|---|---|
| *GBA1* | p.Leu483Pro | 1:155205043-T-C |
| *GBA1* | p.Ser235Pro | Variant49129 |
| *LRRK2* | p.Ile1122Val | 12:40692927-A-G |
| *LRRK2* | p.Ile1122Val | Seq_rs34805604 |
| *LRRK2* | p.Ile2020Thr | 12:40734206-T-C |
| *LRRK2* | p.Ile2020Thr | Seq_rs35870237 |
| *LRRK2* | p.Asn1437Asp | 12:40703027-A-C |
| *LRRK2* | p.Asn1437Asp | Seq_rs74163686 |
| *LRRK2* | p.Arg1441Cys | Seq_rs33939927.1_ilmnfwd_ilmnF2BT |
| *LRRK2* | p.Arg1441Cys | Seq_rs33939927.2_ilmnfwd_ilmnF2BT |
| *LRRK2* | p.Arg1441Cys | Seq_rs33939927.3_ilmnfwd_ilmnF2BT |
| *LRRK2* | p.Arg1441Cys | var_12_40704236 |
| *PINK1* | p.Leu347Pro | 1:20972133-T-C |
| *PINK1* | p.Leu347Pro | Seq_rs28940285 |
| *PINK1* | p.Leu347Pro | rs28940285 |
| *PRKN* | p.Gln34ArgfsTer5 | Variant17166 |
| *PRKN* | p.Cys431Phe | 6:161771237-G-T |
| *PRKN* | p.Glu395Ter | PARK2:NM_004562.2:c.1183G>T:p.(Glu395*) |
| *PRKN* | p.Arg42Pro | 1:35250488-G-C |

| | | |
|---|---|---|
| *SNCA* | p.Ala30Pro | rs104893878 |
| *SNCA* | p.Ala53Thr | seq-rs104893877-B1 |
| *SNCA* | p.Glu46Lys | 4:90749321-G-A |

A table listing additional variants of interest and corresponding Illumina NeuroBooster Array (NBA) probe names. These variants were not identified in short-read Genome Sequencing, hence probe performance could not be assessed.

#### eFigures

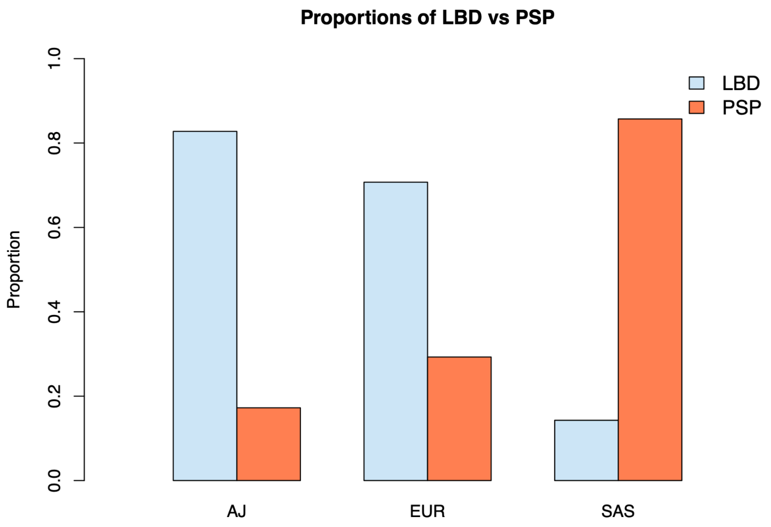

##### eFigure 1. Proportion of LBD and PSP diagnoses across ancestry groups.

Bar plot showing the relative proportion of Lewy body disease (LBD) and progressive supranuclear palsy (PSP) in individuals of Ashkenazi Jewish (AJ), European (EUR) and South Asian (SAS) ancestry. Mutation carriers were excluded from this analysis to avoid confounding effects from *GBA1* and *LRRK2*, which are known to be enriched in AJ. PSP was more frequent in SAS compared with EUR and AJ, while LBD was more frequent in AJ ($p < 0.0001$).

**References**

1. Malek N, Weil RS, Bresner C, et al. Features of GBA-associated Parkinson's disease at presentation in the UK Tracking Parkinson's study. *J Neurol Neurosurg Psychiatry*. 2018;89(7):702-709.

2. Menon PJ, Sambin S, Criniere-Boizet B, et al. Genotype-phenotype correlation in PRKN-associated Parkinson's disease. *NPJ Parkinsons Dis*. 2024;10(1):72.

3. Morales-Briceno H, Ong TL, Duma SR, et al. Recurrent biallelic p.L347P PINK1 variant in Polynesians with parkinsonism and isolated dopa-responsive dystonia. *Mov Disord Clin Pract*. 2022;9(5):696-697.

4. Chen YP, Gu XJ, Ou RW, et al. Genetic analysis of prosaposin, the lysosomal storage disorder gene in Parkinson's disease. *Mol Neurobiol*. 2021;58(4):1583-1592.

5. Beach TG, Adler CH, Sue LI, et al. Arizona Study of Aging and Neurodegenerative Disorders and brain and Body Donation Program. *Neuropathology*. 2015;35(4):354-389.

6. Alafuzoff I, Ince PG, Arzberger T, et al. Staging/typing of Lewy body related α-synuclein pathology: a study of the BrainNet Europe Consortium. *Acta Neuropathol*. 2009;117(6):635-652.
